# High-dose Polyphenol-rich Nutrition Improves Lipid and Inflammation Profiles and Triggers Apoptotic Signaling in Healthy Elderly People (the ErdBEHR Study)

**DOI:** 10.1101/2023.04.03.23287994

**Authors:** Alexander Hartmann, Riccardo Secci, Juliane Mensch, Kathrin Jäger, Elisabeth Steinhagen-Thiessen, Israel Barrantes, Daniel Palmer, Axel Kowald, Rico Schwarz, Burkhard Hinz, Daniela Weber, Tilman Grune, Patricia Hübbe, Gerald Rimbach, Henrik Rudolf, Michael Walter, Georg Fuellen

## Abstract

Nutritional interventions in healthy individuals may be particularly informative if high, but not excessive, amounts of specific healthy foods are taken to maximize effects without sacrificing safety. We hypothesized that high amounts of polyphenols taken on single days may eliminate senescent blood cells. We conducted a ten-week parallel-group controlled randomized open trial with an escalation of consumption, up to ∼4kg fresh strawberries weekly, plus 200g dried strawberries and 240g capers in olive oil on three single “seno-intervention” days, in 168 healthy elderly people aged 50-80 years. Two primary endpoints, LDL cholesterol and high sensitive CRP, were prespecified. We found a significant decline in LDL cholesterol, and in CRP by ∼50% in all groups with seno-intervention days (limited to participants with increased baseline values). LDL levels were reduced by 0.0174 mmol/L for any single 500g-increment in the weekly fresh strawberry intake of the average participant. Gene expression analyses of whole blood suggested improvement of mitochondrial and immunological function, suppression of inflammation (in high-intervention groups), and positive regulation of apoptotic signaling (in the highest-intervention group). Overall, a medium-term nutritional intervention improved lipid and inflammation status, and provided specific hints for apoptotic/senolytic effects.

**Key findings:**

- High-dose polyphenol-rich nutrition improves LDL and CRP levels in healthy elderly
- At highest dose, gene expression data highlight “positive regulation of apoptotic signaling”
- A clear dose-response pattern based on an escalating intervention design

## Introduction

Can food be a preventive medicine and a key to longevity? More specifically, are there particular foods or food combinations and corresponding dosing schemes that provide, on the one hand, both outstanding safety and affordability, and yet strong health benefits on the other? Can aging-associated processes be slowed down considerably, stopped or even partially reversed in humans by dietary geroprotection, that is, by food containing ingredients that promote health? The nutritional “ErdBEHR” (“<u>Erd</u>beeren [strawberries] for <u>B</u>iomarker identification for the <u>E</u>xtension of <u>H</u>ealth by <u>R</u>ejuvenation”) trial reported here was designed to approach these questions for polyphenol-rich food, for which some observational and interventional evidence of positive health effects has been described. Polyphenols are natural organic compounds; frequently studied polyphenols are, e.g., catechins, resveratrol and quercetin, which are found in a variety of plant-based foods. In particular, epidemiologically, health improvements by the Mediterranean and DASH diets are attributed in part to their high polyphenol content (Esposito et al., 2022). Moreover, short- and long-term consumption of polyphenol-rich berries such as strawberries have been associated with a variety of health improvements in previous human studies, with a focus on cardiovascular disease (Secci et al., 2021).

Here, we studied a food-based high-polyphenol intervention based on strawberries, including capers in olive oil in the highest-intervention group. Previously, strawberries were found to contain the polyphenol fisetin that extends healthspan and lifespan in mice (Yousefzadeh et al., 2018), and capers include the closely related polyphenol quercetin (Haytowitz, Wu, & Bhagwat, 2018). Both polyphenols are frequently mentioned in the literature on interventions aiming at removing senescent cells (Wissler Gerdes, Misra, Netto, Tchkonia, & Kirkland, 2021). As described in Box 1, we found ample amounts of bioavailable quercetin in both strawberries and capers. Further bioactive ingredients in strawberries are other polyphenols such as pelargonidin (an anthocyanin responsible for red color), fibers, vitamins and phytosterols (Basu, Nguyen, Betts, & Lyons, 2014). Further, olive oil is also known for its health benefits on lipid metabolism and cardiovascular function and for reducing mortality, due in part to the polyphenols hydroxytyrosol and oleuropein (Guasch-Ferre et al., 2022).

We had three hypotheses: 1) an intervention with fresh strawberries would trigger improvements in the overall cardiovascular risk profile, specifically cholesterol and inflammation status; 2) the intervention would be further ameliorated by adding “seno-intervention days” (SIDs), on 3 single days over the entire 10-week study participation, whereby large but not excessive amounts of freeze-dried strawberries (and, in the highest-intervention group, also capers in olive oil) are consumed; 3) SIDs would trigger the apoptosis of senescent blood cells, leading to hormetic improvements by eliminating these cells. These improvements could thus be triggered by the repair reaction of the body based on the effects of the intervention, resulting in benefits as long as the intervention dose is not too high.

As primary endpoints, we prespecified LDL cholesterol and hsCRP in the ethics application (see Supplementary Files), based on our literature survey (Secci et al., 2021) and a pilot study (see below). We also measured a set of secondary and exploratory endpoints, including gene expression by next-generation-sequencing (transcriptomics). We observed no safety issues except self-reported potential allergy in two cases. We met the primary LDL endpoint in the highest intervention group, culminating a dose-response trend over all intervention groups, which was even stronger for total cholesterol. We found anti-inflammatory effects in the high-intervention groups, superimposed by a pro-inflammatory pattern in the highest-intervention group, just after the last SID, which we attribute to apoptotic processes based on the gene expression data. Hypothetically, the highest-intervention SID enabled the killing of senescent blood cells, an exciting proposition deserving further research. Since polyphenols may interact with medication (Lopes, Coimbra, Costa, & Ramos, 2021), (Lopez-Yerena, Perez, Vallverdu-Queralt, & Escribano-Ferrer, 2020) (Box 2), the study was performed with healthy individuals.

## Methods

We follow the Consort reporting guidelines for describing the study.

### Trial Design

Overall, we adopted a parallel-group 5-arm single-center controlled randomized and open design (Fig. 1). The control group (group 1) featured a small amount of strawberry consumption (once a week), and the four intervention groups (groups 2-5) featured a longitudinal escalation over the 10-week participation period. Three intervention groups also included three “seno-intervention days” (SIDs) each, with an escalating dose of freeze-dried strawberries, adding capers in olive oil in group 5. Four visits to the study center were done over the 10 weeks of study participation; at the start (t1); after two weeks (day 14, t2), after another 7 weeks (day 63, t3) and at the end (day 70, t4).

**Figure 1.**
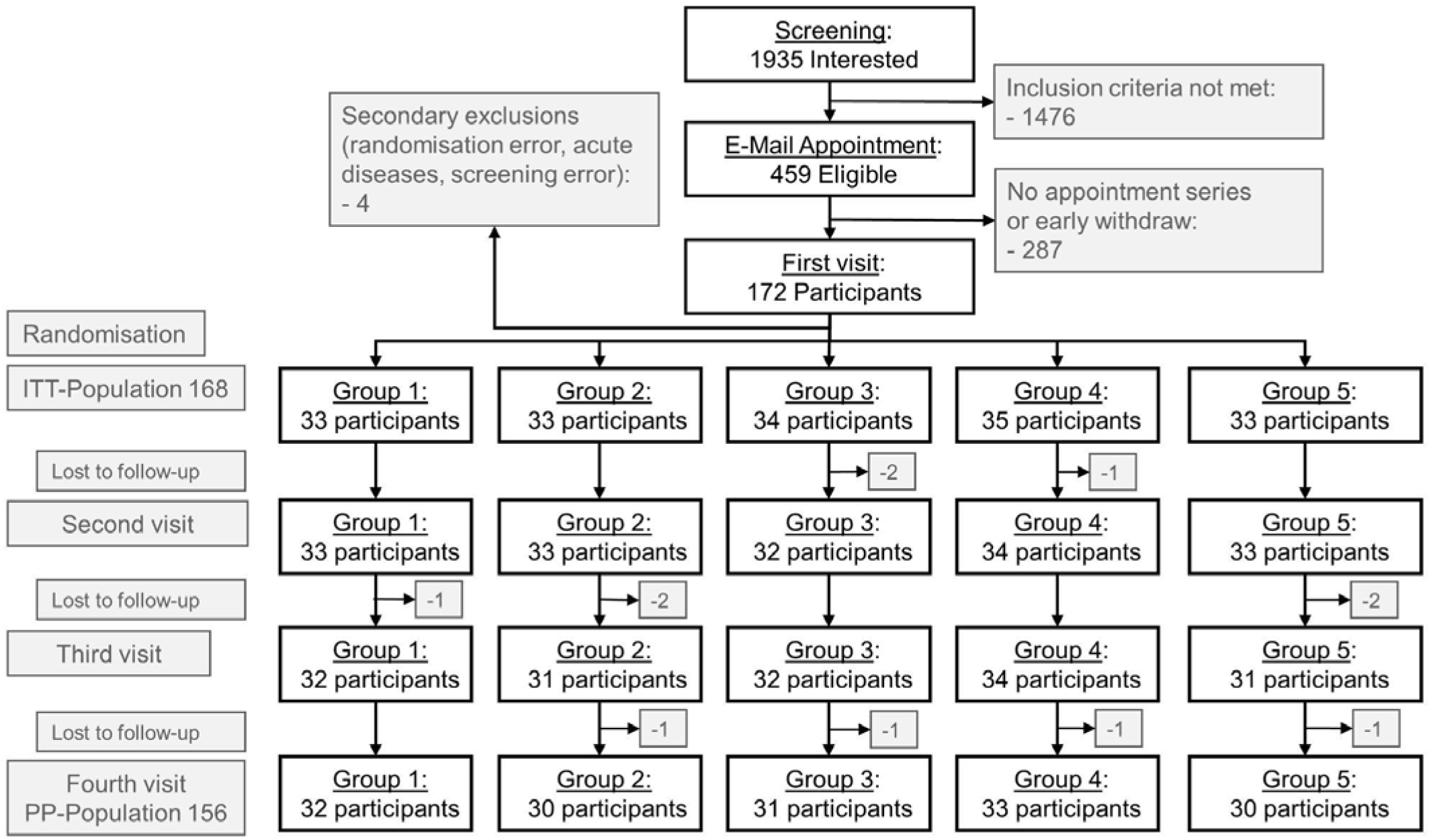
Flowchart of the ErdBEHR study. Abbreviations: ITT Intention to treat, PP per-protocol.

### Participants

Based on a pilot trial in August 2020, the study was powered as described below. Recruitment of participants was conducted via a recruitment questionnaire run in early 2021 (see Supplementary Files (listing on page 20), in German) written by the investigators but run and advertised by the supporting strawberry farm, Karls Erdbeerhof, Rövershagen, Germany. Based on the predefined inclusion and exclusion criteria as outlined in the Ethics Application (see Supplementary Files, in German), respondents qualified if they confirmed the following self-reported attributes at the time of their response.

- No prescription drugs (except cholesterol-lowering drugs and antihypertensive drugs; thyroid medication was allowed ad-hoc) and no severe chronic diseases.
- No known food allergy/intolerance, no food allergy/intolerance to strawberries in direct relatives.
- Strawberry salespoint of Karls Erdbeerhof easily accessible, to obtain fresh fruit regularly.
- No intake of dietary supplements or vitamin supplements exceeding standard recommendations.
- Consumption of the intervention food in the past, that is, weekly in the summer for strawberries; weekly, monthly or rarely for capers.
- Weight <= 100kg (for reasons of “dosage” of the intervention food).
- Age 50-80 years at time of responding to the questionnaire.

### Data Collection

Visits to the study center were carried out Tuesdays to Fridays 7-10h (avoiding Mondays to mitigate weekend effects), on participation days 1, 14, 63 and 70 of the study. Participants were asked to fast for at least 8 hours (only water and medications as noted above were allowed). Great care was taken to minimize deviations from the timeplan. Aside from the blood draws, blood pressure, pulse, and a Chair-Rise-Test were also performed. Participants were asked about their adherence on days 14, 63 and 70, and protocol violations were recorded. Almost all participants filled in the validated EPIC-Potsdam Food Frequency Questionnaire-II (Nothlings, Hoffmann, Bergmann, & Boeing, 2007), (Freese et al., 2014) within the first weeks of the study. Clinical data questionnaires (see Supplementary Files, in German) were given to participants on days 1 and 63 of the study, to be returned on days 14 and 70, including quality-of-life questionnaires (EQ-5D-5L and EQ-VAS), a fatigue questionnaire, self-reported anthropometrics, current disease and medication, COVID-19 disease and/or vaccination, vitamin and supplement consumption (regular / in excess of suggested intake), smoking (in pack-years), and exercise. On day 1, history of disease and direct relatives’ disease was also tallied. Questionnaire data were digitized following the 4-eyes principle.

Up to 63mL of blood were drawn and used to measure standard parameters, including platelets, neutrophils, lymphocytes (giving rise to neutrophil-lymphocyte-ratio), as well as glucose and insulin/C-peptide (giving rise to HOMA-IR), HbA1c, alkaline phosphatase, electrolytes (Calcium, Natrium, Kalium), liver enzymes (e.g., GGT), albumin, creatinine (giving rise to estimated glomerular filtration rate, eGFR), ferritin, coagulation markers (Factor 5, Factor 8, Factor 12 and d-dimers) and gene expression by next-generation sequencing (see below). Moreover, Luminex protein data of IL6, sTNFr1 and GDF15 were obtained from the serum of 143 compliant non-smoking participants (by whom no difficulties in food consumption were reported in the questionnaires), by using premixed multiplex Human magnetic Luminex^®^ Assay plates (R&D Systems/Bio-Techne, Minneapolis USA) analyzed on a Luminex^®^ 100/200^TM^ (Burlington USA). Vitamins and carotenoids (retinol, α-carotene, β-carotene, α-tocopherol, γ-tocopherol, lutein, lycopin, β-cryptoxanthin) were analyzed in plasma by reversed-phase HPLC with UV detection (internal standard and carotenoids) and fluorescence detection (retinol and tocopherols), respectively (Stuetz et al., 2016). MDA (malondialdehyde) was measured by reverse-phase HPLC and fluorescence detection after derivatization with thiobarbituric acid (Weber et al., 2017). Lipidomics, proteomics, metabolomics, and analyses of the optional skin biopsies and stool microbiomes are pending.

### Interventions

The control group 1 was instructed to eat 500g of fresh strawberries once a week. A placebo was not available, as a convincing placebo for whole, fresh strawberries is not available/practical. 500g of strawberries per week matches the moderate consumption reported in the recruitment questionnaire by all qualifying respondents. Our design further followed a “double-escalation” strategy: the total polyphenol consumption was increased from group to group by introducing specific dietary components; at the same time, all groups except for the control group featured an escalation of dose in time. The standard intervention group 2 was thus instructed to eat an escalating dose of fresh strawberries, 3 times a week 500g over the first two weeks, 5 times 500g each week over the next 7 weeks, and 5 times 750g over the last week. The high-intervention groups all featured an additional three SIDs, of escalating dose. The first SID was scheduled for day 1, *after* the t1 blood draw; with the second and the third SIDs scheduled for the day *preceding* the t2 and the t4 blood draws, respectively. In group 3, the SIDs consisted of 100g dried strawberries; in group 4, the first SID featured 100g, but the second and third featured 200g of dried strawberries; in group 5, an additional 120g of capers in olive oil were added on the first SID, and 240g on the second and third SID. Participants were explicitly allowed to distribute the SID consumption over the two days before the blood draw, if, based on their experiences on day 1, they felt that they could not consume all SID food on a single day.

Fresh strawberries were provided for free by the supporting strawberry farm Karls Erdbeerhof; participants received vouchers which they were asked to redeem at a salespoint they could reach easily and they were told that freshness of the berries is important. Freeze-dried strawberries and capers in “native olive oil extra” (Rapunzel, Legau, Germany) were provided by *Karls Erdbeerhof* and handed out at the study center. The variety of fresh strawberry consumed was not recorded. Every two weeks, strawberry samples were frozen for future analyses. Strawberry samples from August 2021 (varieties: *Milwana* and *Florentina*) as well as the capers were subjected to mass-spectrometry analysis of selected polyphenols (fisetin and quercetin), see Box 1. Participants were instructed to adhere to the consumption plan, but to not eat the food if it was causing a high degree of aversion; in this case they were instructed to inform study personnel at their next visit. Further, participants were explicitly allowed to consume other high-quality fresh strawberries according to consumption plan for at most two weeks if they traveled away from a salespoint of Karls Erdbeerhof. The instructions also included explicit statements to not change dietary habits otherwise; to not destroy the health-promoting ingredients of the intervention food e.g. by cooking or baking; to consume the olive oil together with the capers (if any); to not consume other food with a specific health-related claim (mentioning green tea or garlic as examples) regularly and in high amounts, and they were given a sheet with 5 recipes compatible with these guidelines (see Supplementary Files, in German) as well as detailed personalized consumption plans (for a sample, see Supplementary Files, in German).

### Outcomes

In the March 2021 ethics application (see Supplementary Files, in German), we prespecified LDL and hsCRP as primary endpoints, and the following secondary endpoints: Chair-Rise-Test (number of cycles of rising and sitting accomplished in 30 seconds, CR30s), quality-of-life questionnaires (EQ-5D-5L, EQ-VAS, collectively called EQs), HDL cholesterol, total cholesterol (TC), triglycerides (TG), HOMA-IR, *Phenotypic Age* (Levine, 2013), IL6, sTNFr1 and GDF15. The primary endpoints were motivated by (Secci et al., 2021) and the pilot study. Our assessment of the literature further motivated the lipid and glucose metabolism markers (Daneshzad, Shab-Bidar, Mohammadpour, & Djafarian, 2019), (Fallah, Sarmast, & Jafari, 2020; Shah & Shah, 2018), the IL6 and sTNFr1 combination (Justice et al., 2018), and GDF15 (Schafer et al., 2020; Tanaka et al., 2018). *Phenotypic Age*, CR30s and EQs were recommended in expert interviews.

### Sample size, Randomization and Blinding

In a 19-participant 4-group unpublished pilot trial in August 2020, we had found that one week of fresh strawberry consumption and two SIDs one week apart were associated with improvements in LDL cholesterol and CRP. Based on the results of the pilot study, sample size was planned using the R-package *WebPower* for repeated measures ANOVA, assuming a between-groups effect size of 0.4 and sphericity of at least 0.6, which gave a sample size of 180 participants for a power of 80% to detect a group-by-time interaction, reflecting the hypothesis of differences in change over time between groups. The study was planned for two primary endpoints, namely LDL and hsCRP, applying a type I error of 2.5%. The randomization list was generated with a block size of 10 to avoid imbalances of allocation within weeks, and with stratification by smoking status only. Investigators were blinded; allocation to groups was not revealed to the physicians or laboratory personnel involved. Participants were not blinded to groups.

### Statistical methods

Participant characteristics at baseline were tabled by group assignment with mean (and standard deviations) or count (and percentage). Since repeated measures ANOVA invokes risk of bias in estimates when used with drop-outs, statistical analyses were done by means of linear mixed models (LMM) (Cnaan, Laird, & Slasor, 1997) for all outcomes unless noted otherwise. CRP values were log-transformed, and for Chair-Rise-Test a generalized model for poisson data was applied. Primary endpoints were examined by LMM, reporting statistical test results for group-by-visit interaction (i.e., for differences in change over time between groups), estimated marginal means (EMM) for improvements in outcomes (from t1 to t4) for each group, and statistical test results for treatment effects, as the difference in improvement, of groups 2-5 versus the control group 1. Secondary endpoints were also tested in LMMs (unless stated otherwise), checking group-by-visit interactions first. If these interactions may have triggered a notable difference in time trends between groups, which we deemed possible in case of *p<0.2*, effects in groups were examined separately, otherwise the change in comparison to baseline irrespective of group (i.e., the effect of time considering the fourth visit) was reported. Confirmatory, explorative and descriptive analyses were performed with the statistical software R, along with the R-packages lmerTest, emmeans, comparegroups, and ggplot. The significance level was set to 0.025 for the two primary endpoints, and 0.05 otherwise, and all *p-* values are two-sided.

### Sequencing and bioinformatics methods

We profiled 36 compliant nonsmoking participants by next-generation sequencing of PBMC at timepoints t1 and t4, that is, 5 participants each randomly selected from groups 1, 2 and 4, and all 21 compliant nonsmoking participants from group 5. RNA quantity and purity was measured by determining the UV/VIS absorbance (NanoDrop One, Thermo Fisher Scientific) at three analytical wavelengths: *230, 260* and *280 nm*. 3’mRNA sequencing libraries were then generated using the QuantSeq 3’mRNA-Seq Library Prep Kit (Moll, Ante, Seitz, & Reda, 2014) according to manufacturer’s instructions (Lexogen, Austria), and sequenced on a HiSeq4000 platform (Illumina, San Diego) using a single-end protocol with an RNA read-length of 100, at the Cologne Center for Genomics (University of Cologne, Germany). The gene expression analysis of the samples started with Salmon version 1.9.0 (Patro, Duggal, Love, Irizarry, & Kingsford, 2017), mapping the RNA reads to the human transcriptome GRCh38, as retrieved from Ensembl. For each sample, the two technical replicates were combined by summing the gene counts. DESeq2 release 3.16 (Love, Huber, & Anders, 2014) was then used to calculate differential gene expression (log fold change, p-value) between timepoints, for each group. A PCA of the samples was created, using the R function plotPCA from the R package BiocGenerics version 0.42.0 (Huber et al., 2015), using the BiocGenerics parameter *ntop = 500* to focus on the top 500 variable genes across the samples, which are more likely to be differentially expressed. The function *gconvert* was used to convert the gene IDs from the Ensembl format to the Entrez_gene format. An enrichment analysis was then carried out with GSEA version 4.3.1 (Subramanian et al., 2005) using the R library clusterProfiler, for the Gene Ontology (GO) and KEGG. We only report significant enrichments (with a negative log-normalized *q-value* ≥ *1.5*). REVIGO version 2015-02-17 (Supek, Bosnjak, Skunca, & Smuc, 2011) was used to reduce the number of GO terms by half (reduction parameter set to 0.5, using *homo sapiens* as *species* parameter and *higher absolute value* as *Enrichment score* parameter). Barplots were created to show the terms featuring the at most 25 terms with highest absolute *Enrichment scores*. Heatmaps were prepared using the R package Pheatmap.

### Ethics

The ethics board of the Rostock University Medical Center approved this study under the registration number A 2021-0096 (see Supplementary Files, in German). The study was registered as <u>DRKS00026998</u> at the German register for clinical trials (DRKS). We confirm that the study conforms to recognized standards and that participants have given free prior informed consent.

## Results

### Participant Flow

Via the recruitment questionnaire (see Supplementary Files, in German), 1935 applications were received and screened (Figure 1) in early 2021. According to the inclusion/exclusion criteria, 459 applicants (including smokers) qualified, who were invited consecutively. Series with four appointments per applicant on participation days 1, 14, 63 and 70 were arranged by email. After informed consent, 172 participants could be allocated to one of the 180 appointments series (of 4 visits each) that were available, from June 10 to October 1, 2021, recruiting ∼28 participants per week from June 10 to July 23, 2021. After baseline assessment, participants received bags packed according to group allocation, containing vouchers for fresh strawberries and the personalized consumption plan, as well as (for groups 3-5) the additional SID foods to be consumed until the next appointment. There were four secondary exclusions due to randomization error (1), acute disease/accident (2), and screening error (1) leaving 168 participants for analysis in the *intention to treat* (ITT) population. 12 participants were lost to follow-up, so that 156 finished the study regularly.

### Baseline Data

The baseline characteristics of the 168 participants are shown in Table 1. Average age was ∼60 years, 66% were female. Participants’ characteristics were balanced between groups for the clinical attributes and biomarkers at baseline, with the exception of age which had a slight random imbalance.

**Table 1:** Baseline data of the ErdBEHR study. Baseline characteristics, and selected primary and derived biomarkers of participants; laboratory biomarkers are listed alphabetically. Numbers show mean (with standard deviation), or count (with percentage) according to type of characteristic/biomarker. Abbreviations: BMI: body-mass index; CRP: C-reactive Protein; GGT: gamma-glutamyltransferase; HbA1c: glycated hemoglobin A1c; HDL: high-density lipoproteins cholesterol; LDL: low-density lipoproteins cholesterol; HOMA-IR: Homeostatic Model Assessment for Insulin Resistance; eGFR: estimated glomerular filtration rate, PhenoAge: phenotypic age (by Levine).

|  |  | Group 1 | Group 2 | Group 3 | Group 4 | Group 5 | *p-value* |
| --- | --- | --- | --- | --- | --- | --- | --- |
|  | **N=168** | **N=33** | **N=33** | **N=34** | **N=35** | **N=33** |  |
| Age | 60.7<br>(7.32) | 57.8<br>(5.66) | 61.9<br>(8.14) | 60.1<br>(6.69) | 60.4<br>(8.15) | 63.0<br>(6.97) | 0.044 |
| Sex (female) | 110<br>(65.5%) | 20<br>(60.6%) | 25<br>(75.8%) | 25<br>(73.5%) | 20<br>(57.1%) | 20<br>(60.6%) | 0.369 |
| Smoking (yes) | 16<br>(9.52%) | 3<br>(9.09%) | 3<br>(9.09%) | 3<br>(8.82%) | 4<br>(11.4%) | 3<br>(9.09%) | 1.000 |
| BMI | 25.9<br>(3.49) | 26.1<br>(2.96) | 25.8<br>(3.41) | 25.4<br>(4.20) | 25.7<br>(2.90) | 26.8<br>(3.86) | 0.585 |
| Weight (kg) | 75.8<br>(12.1) | 77.8<br>(9.85) | 73.7<br>(11.6) | 72.6<br>(14.7) | 77.8<br>(10.5) | 77.2<br>(13.1) | 0.227 |
| Albumin (g/L) | 43.4<br>(2.02) | 43.4<br>(1.62) | 43.3<br>(2.11) | 43.3<br>(1.98) | 43.4<br>(2.11) | 43.8<br>(2.32) | 0.883 |
| Alkaline phosphatase (U/L) | 76.4<br>(20.1) | 73.6<br>(17.5) | 79.0<br>(21.7) | 76.8<br>(22.2) | 77.0<br>(19.8) | 75.4<br>(19.5) | 0.866 |
| Total Cholesterol (mmol/L) | 6.04<br>(1.10) | 5.92<br>(1.16) | 6.13<br>(1.18) | 6.01<br>(1.07) | 6.11<br>(1.08) | 6.03<br>(1.08) | 0.941 |
| Creatinine (μmol/L) | 73.3<br>(13.1) | 71.1<br>(12.3) | 72.5<br>(15.4) | 73.1<br>(13.6) | 76.0<br>(13.3) | 73.6<br>(10.7) | 0.629 |
| CRP (mg/L) | 1.87<br>(1.51) | 1.91<br>(1.81) | 1.96<br>(1.49) | 1.56<br>(0.94) | 1.76<br>(1.31) | 2.20<br>(1.85) | 0.499 |
| Ferritin (μg/L) | 156<br>(138) | 162<br>(125) | 134<br>(128) | 146<br>(130) | 148<br>(93.5) | 195<br>(197) | 0.461 |
| GGT (U/L) | 27.2<br>(24.9) | 21.1<br>(11.6) | 29.7<br>(23.6) | 24.8<br>(15.4) | 25.4<br>(18.9) | 35.1<br>(42.3) | 0.189 |
| Glucose (mmol/L) | 5.71<br>(0.67) | 5.84<br>(1.00) | 5.69<br>(0.50) | 5.76<br>(0.56) | 5.56<br>(0.44) | 5.72<br>(0.70) | 0.535 |
| HbA1c (mmol/mol Hb) | 39.3<br>(4.31) | 39.6<br>(7.13) | 38.7<br>(3.11) | 40.8<br>(3.21) | 38.6<br>(3.34) | 39.0<br>(3.18) | 0.220 |
| HDL (mmol/L) | 1.72<br>(0.42) | 1.64<br>(0.34) | 1.68<br>(0.41) | 1.88<br>(0.46) | 1.69<br>(0.48) | 1.72<br>(0.41) | 0.175 |
| Interleukin 6 (pg/mL) | 3.02<br>(1.43) | 3.03<br>(1.43) | 3.17<br>(1.44) | 3.07<br>(1.51) | 3.15<br>(1.41) | 2.67<br>(1.33) | 0.380 |
| LDL (mmol/L) | 3.86<br>(0.89) | 3.85<br>(0.91) | 3.98<br>(0.96) | 3.69<br>(0.87) | 3.96<br>(0.85) | 3.82<br>(0.87) | 0.683 |
| Neutrophil-lymphocyte-ratio | 1.80<br>(0.73) | 1.73<br>(0.60) | 1.95<br>(0.85) | 1.61<br>(0.58) | 1.92<br>(0.87) | 1.78<br>(0.69) | 0.282 |
| Platelets (10E9/L) | 240<br>(54.3) | 246<br>(57.8) | 227<br>(54.3) | 239<br>(36.8) | 253<br>(57.8) | 235<br>(61.4) | 0.342 |
| Triglycerides (mmol/L) | 1.16<br>(0.55) | 1.19<br>(0.42) | 1.07<br>(0.45) | 1.13<br>(0.64) | 1.18<br>(0.61) | 1.25<br>(0.62) | 0.765 |
| HOMA-IR [mmol/L] | 0.20<br>(0.09) | 0.21<br>(0.11) | 0.20<br>(0.08) | 0.20<br>(0.09) | 0.18<br>(0.06) | 0.20<br>(0.09) | 0.715 |
| eGFR [mL/min] | 80.3<br>(13.8) | 85.0<br>(13.8) | 79.4<br>(15.5) | 78.7<br>(13.6) | 78.9<br>(13.8) | 79.7<br>(12.1) | 0.305 |
| PhenoAge [years] | 54.9<br>(8.36) | 52.2<br>(6.81) | 56.3<br>(9.12) | 54.3<br>(7.90) | 54.9<br>(9.27) | 56.8<br>(8.14) | 0.219 |
| Health (EQ-5d-5L) | 0.94<br>(0.10) | 0.95<br>(0.07) | 0.95<br>(0.06) | 0.91<br>(0.16) | 0.94<br>(0.11) | 0.94<br>(0.06) | 0.362 |
| Health (EQ-VAS) | 82.5<br>(14.6) | 83.5<br>(10.6) | 83.74<br>(10.6) | 81.0<br>(13.4) | 83.7<br>(12.1) | 85.9<br>(9.64) | 0.287 |
| 30s-Chair-Stand-Test | 13.2<br>(2.53) | 13.8<br>(2.14) | 12.7<br>(2.11) | 13.5<br>(2.53) | 13.6<br>(2.88) | 12.7<br>(2.80) | 0.241 |

### Prespecified primary and secondary endpoints: Dietary intervention improves LDL, CRP and Total Cholesterol in healthy elderly

As the first primary endpoint, we pre-specified LDL in the March 2021 ethics application (see Supplementary Files, in German), and we noted a significant decline in LDL cholesterol after ten weeks in group 5 versus group 1 (–0.148 mmol/L, that is, –5.72 mg/dL, *p=0.007*), see Figure 2a and Table 2. We also noted a lowering of LDL values for all other intervention groups. As the second primary endpoint, we pre-specified hsCRP. We found no significant intervention effects in the hsCRP data as recorded (Table 2); we found that these were measured with a detection limit of 1 mg/L, triggering a floor effect (40% of participants had a CRP baseline value recorded at the detection limit). Intervention effects for the subgroup with elevated starting values (>2 mg/L), justifying the use of statins in primary prevention (Kaptoge et al., 2010), (Buckley, Fu, Freeman, Rogers, & Helfand, 2009), (Helfand et al., 2009), are clinically most meaningful. Data of the 43 qualifying participants were thus analyzed exploratively, henceforth referred to as CRP data. While the change over time between groups overall was insignificant, groups 3, 4 and 5 featuring SIDs had significantly lower CRP levels compared to groups 1 and 2, see Figure 2b and Table 2, even though CRP values are expected to increase over summer (see (Dopico et al., 2015), their fig. 5d).

**Figure 2.**
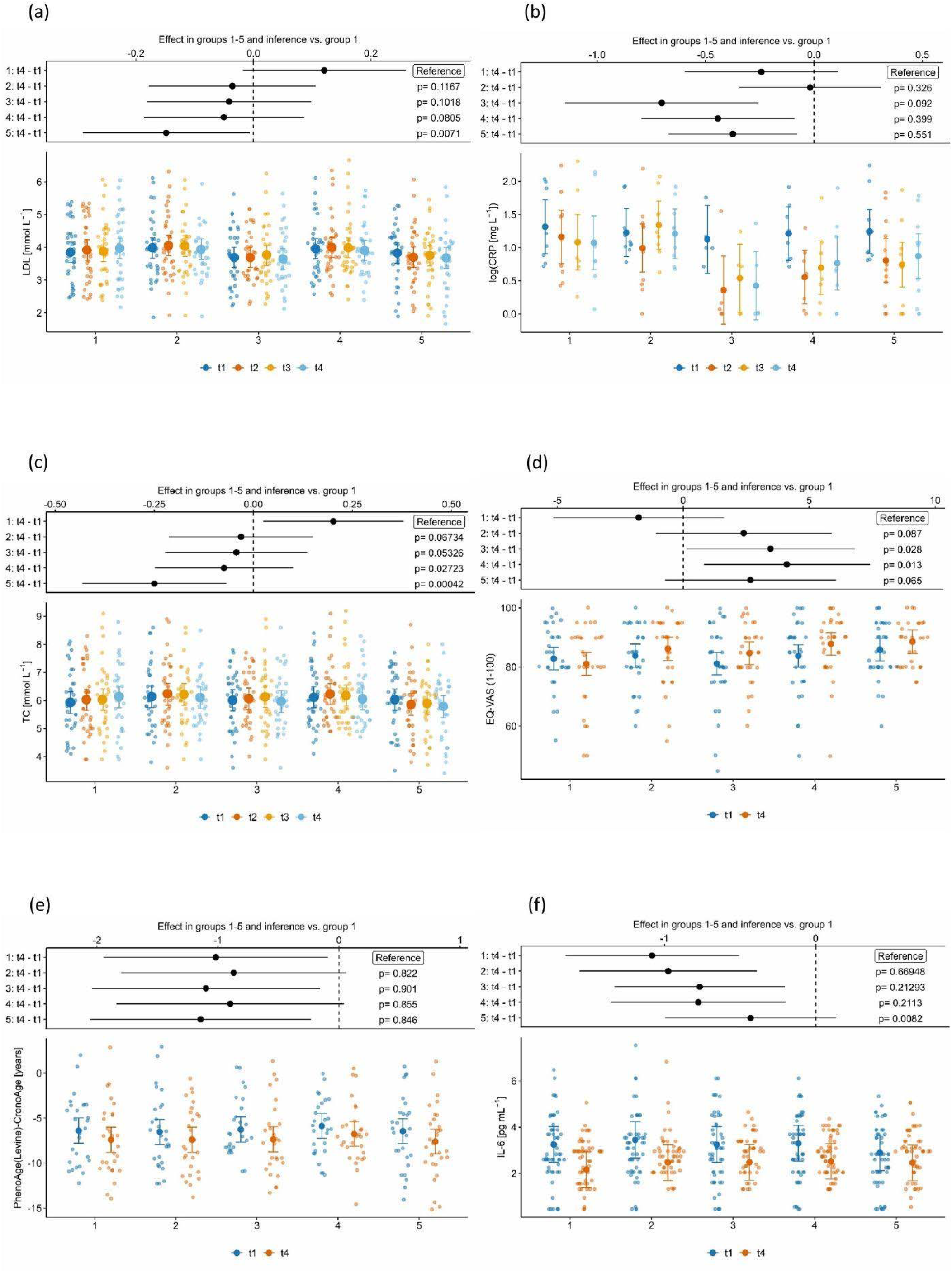
Effect plots and response data plots for primary and selected secondary endpoints. (a) LDL; (b) CRP of participants with high baseline level; (c) total cholesterol; (d) EQ-VAS; (e) *Phenotypic Age*; (f) IL6. Effects in groups (top) are the differences between last visit (t4) and first visit (t1) of estimated marginal means (EMM) with 95% confidence intervals from the linear mixed model. Dots in plots (bottom) represent model-based individual response data across groups and timepoints (t1, t2; t3, t4 as available), including their EMM (visualized by a large dot) and confidence interval.

**Table 2:** Effect estimates and statistical tests for primary, secondary and selected exploratory outcomes. Results for primary, secondary and selected exploratory analyses. Numbers show effect estimates (in units as in Table 1, except for logarithmic transformation) with standard errors and p-value according to chisquare-test or t-test as specified in the Methods section. Abbreviations: See Table 1; ITT, intention to treat analysis. * Detection limit of 1 mg/dL, values below 1 were not set to missing, instead the value 1 was inserted for ITT analysis of group-by-time interaction. ** Same for all groups.

| Outcome<br>(population, *n*) | Kind of effect (interaction effect, or effect on outcome) being tested/<br>estimated | Effect estimates;<br>change at last visit vs first visit<br>(standard error) | *p*-value |
| --- | --- | --- | --- |
| LDL<br>(ITT, *n*=168) | Group-by-visit interaction |  | 0.248 |
|  | Effect by group at last visit |  |  |
|  | group 1 | 0.121 (0.069) | (Ref.) |
|  | group 2 | -0.036 (0.071) | 0.117 |
|  | group 3 | -0.041 (0.070) | 0.101 |
|  | group 4 | -0.050 (0.068) | 0.081 |
|  | group 5 | -0.148 (0.071) | 0.007 |
| CRP (log-transf.)<br>(ITT, *n*=168)* | Group-by-visit Interaction |  | 0.337 |
| CRP (log-transf.)<br>(elevated at baseline: >2 mg/L,<br>*n*=43) | Group-by-visit Interaction |  | 0.100 |
|  | Effect by group at last visit |  |  |
|  | group 1 | -0.243 (0.179) | (Ref.) |
|  | group 2 | -0.018 (0.165) | 0.336 |
|  | group 3 | -0.702 (0.226) | 0.092 |
|  | group 4 | -0.443 (0.179) | 0.399 |
|  | group 5 | -0.374 (0.150) | 0.551 |
| TC<br>(*n*=168) | Group-by-visit Interaction |  | 0.091 |
|  | Effect by group at last visit |  |  |
|  | group 1 | 0.202 (0.090) | (Ref.) |
|  | group 2 | -0.031 (0.092) | 0.072 |
|  | group 3 | -0.043 (0.091) | 0.057 |
|  | group 4 | -0.074 (0.089) | 0.030 |
|  | group 5 | -0.250 (0.092) | <0.001 |
| EQ-5D-5L<br>(*n*=164) | Group-by-visit Interaction |  | 0.731 |
|  | Effect at last visit** | -0.005 (0.009) | 0.599 |
| EQ-VAS<br>(*n*=164) | Group-by-visit Interaction |  | 0.115 |
|  | Effect by group at last visit |  |  |
|  | group 1 | -1.8 (1.71) | (Ref.) |
|  | group 2 | 2.4 (1.77) | 0.092 |
|  | group 3 | 3.5 (1.69) | 0.031 |
|  | group 4 | 4.1 (1.67) | 0.015 |
|  | group 5 | 2.7 (1.72) | 0.069 |
| Phenotypic Age<br>(*n*=143) | Group-by-visit Interaction |  | 0.992 |
|  | Effect at last visit** | -1.01 (0.207) | <0.001 |
| HOMA-IR<br>(*n*=156) | Group-by-visit Interaction |  | 0.114 |
|  | Effect by group at last visit |  |  |
|  | group 1 | -0.011 (0.008) | (Ref.) |
|  | group 2 | -0.017 (0.008) | 0.609 |
|  | group 3 | -0.014 (0.008) | 0.817 |
|  | group 4 | 0.010 (0.008) | 0.052 |
|  | group 5 | -0.008 (0.008) | 0.762 |
| Triglycerides<br>(*n*=168) | Group-by-visit Interaction |  | 0.230 |
|  | Effect at last visit** | 0.055 (0.050) | 0.271 |
| HDL cholesterol<br>(*n*=168) | Group-by-visit Interaction |  | 0.882 |
|  | Effect at last visit** | 0.042 (0.013) | 0.001 |
| Interleukin-6<br>(*n*=142) | Group-by-visit Interaction |  | 0.089 |
|  | Effect by group at last visit |  |  |
|  | group 1 | -1.081 (0.281) | (Ref.) |
|  | group 2 | -0.975 (0.288) | 0.669 |
|  | group 3 | -0.766 (0.288) | 0.213 |
|  | group 4 | -0.776 (0.284) | 0.211 |
|  | group 5 | -0.430 (0.278) | 0.008 |
| sTNFr1<br>(*n*=143) | Group-by-visit Interaction |  | 0.472 |
|  | Effect at last visit** | -130.4 (47.95) | 0.008 |
| GDF15<br>(*n*=124) | Group-by-visit Interaction |  | 0.450 |
|  | Effect at last visit** | -47.3 (26.29) | 0.081 |
| Chair-Rise-Test<br>(generalized linear<br>mixed model,<br>Poisson, *n*=156) | Group-by-visit Interaction |  | 0.999 |
|  | Effect at last visit** | 0.16 (0.029) | <0.001 |

Regarding the prespecified secondary endpoints, TC changes over time were different between groups (*p=0.091)*, and groups 4 and 5 improved significantly over group 1 by up to –0.25 mmol/L longitudinally (and by –0.45 mmol/L, group 5 in comparison to group 1), see Figure 2c and Table 2. Quality of life could be evaluated in 152 participants, and for EQ-5D-5L (subjective health in five dimensions of interest), no differences between groups were found. For EQ-VAS (subjective health on a scale from 0 to 100), there was no significant group-by-time interaction (*p=0.115*), but groups 3 and 4 improved significantly versus group 1 by up to 6 points (Figure 2d and Table 2). Regarding the further pre-specified secondary endpoints, there were no groupwise differences in change over time for *Phenotypic Age* (increasing in all groups by one year, *p=0.032*, Figure 2e), HOMA-IR, HDL cholesterol (increasing in all groups by 0.042 mmol/L, *p=0.001,* Suppl. Fig. 1a, at end of manuscript file), triglycerides, IL6 (decreasing in all groups over time, but significantly least so in group 5, Figure 2f and Table 2), sTNFr1 (decrease in all groups, Suppl. Fig. 1b) and GDF15 (decrease in all groups, Suppl. Fig. 1c). Chair rise test data showed that the number of attempts in 30 seconds was improved on average by 2 repetitions, with no differences between groups.

### Exploratory analyses (fatigue, clinical chemistry, micronutrients, MDA, LDL elevated at baseline)

Further testing included other biomarkers that may benefit from strawberry intake according to expert consultations. For eGFR, we found no significant differences in change over time between groups; we found higher values for groups 4 and 5, though (Suppl. Fig. 1d). Other exploratory analyses of fatigue, platelets, neutrophil-lymphocyte-ratio, HbA1c, alkaline phosphatase, electrolytes, liver enzymes, albumin, and ferritin showed no group-specific effects, but platelet counts increased in all groups, see Suppl. Fig. 1e. Furthermore, we found no significant differences between groups for β-carotene (*p=0.086*), even though values in group 2-5 decreased in comparison to group 1 (Suppl. Fig. 1f). There were no specific findings for the other micronutrients, nor for MDA levels, nor for the coagulation markers, i.e., Factor 5, Factor 8, Factor 12 and d-dimers. Exploratively, we also analyzed the subgroup of participants which had high starting values in the primary endpoint LDL, consisting of 72 participants with elevated LDL (>4 mmol/L). Whereas in group 1, LDL continued to rise, groups 2, 4 and 5 significantly improved, on average by –0.2 mmol/L.

### Dose-response analysis of intervention food intake on LDL levels

Food intake was escalated over groups and time. To estimate effects of food doses, we modeled intake in the time period preceding the LDL measurements, and the group variable was decomposed into weekly food intake of fresh strawberries, modeled with unity 500g/week, capers and olive oil (120g/week), and freeze dried strawberries (100g/week). LDL levels were found to be reduced by 0.0174 mmol/L (0.67 mg/dL), for any 500g-increment in the weekly fresh strawberry intake of an average participant (*p=0.011*), corresponding to an expected change of –0.13 mmol/L (–5 mg/dL) for the study maximum intake of up to 5 · 750g weekly (that is, 3750g weekly); there was no clear signal for dried strawberries, nor for capers in olive oil considered alone.

### Gene expression: Dietary intervention suppresses inflammation, but also induces apoptosis and phagocytosis pathways

Enrichments of GO biological processes (GOBP) and KEGG pathways are presented in the following with a focus on the longitudinal changes in the intervention groups. As expected, the PCA (Suppl. Fig. 2) tends to group the samples by participant ID, motivating longitudinal analyses. The Supplementary Data (see listing on page 20), include the tables of DEGs, the enrichments (tables and additional figures), as well as additional results and discussion of the control group 1 gene expression data.

Considering group 2 and group 4, we profiled 5 participants each, longitudinally at the first and the last time point. Inspecting GOBP terms, for group 2 we found a significant positive enrichment (that is, enrichment within the upregulated genes, “enriched-up”) of terms related to mitochondrial respiration (aerobic electron transport chain / mitochondrial respiratory chain complex assembly) and coagulation (platelet activation / coagulation) (Figure 3a). Terms with a significant negative enrichment (that is, enrichment within the downregulated genes, “enriched-down”) pointed to a reduction in inflammation (toll−like receptor signaling pathway / myeloid leukocyte differentiation / (regulation of) interleukin−6 production). Correspondingly, based on KEGG, *oxidative phosphorylation / thermogenesis* and *platelet activation* are “enriched-up” together with cardio-related pathways and general disease pathways (Figure 3b). The latter were enriched because they include the mitochondrial complexes and “housekeeping proteins’’ such as tubulin, actin, ubiquitin and histones. In turn, “enriched-down” pathways are associated with lipids (*fatty acid metabolism*) and inflammation (TNF-, IL-17- and toll-like signaling and inflammatory diseases). Employing GOBP for group 4, we again saw an “enriched-up” of terms related to mitochondrial respiration (mitochondrial respiratory chain complex assembly / mitochondrial ATP synthesis coupled electron transport) and additionally a higher activity in some immune-associated processes (myeloid leukocyte mediated immunity / immune effector process) (Figure 3c). Furthermore, we saw an increase in reactive oxygen species metabolism (matching the increase in mitochondrial respiration), and in metabolic activity, e.g. catabolism of lipids and carbohydrates. Among the terms reflecting downregulation, we found a variety of terms, many of which related to cellular stress. Using KEGG, we also found upregulation of respiration/metabolism (*oxidative phosphorylation / thermogenesis* / *glycolysis/gluconeogenesis),* and related (disease) terms, and we found downregulation related to cancer and (again) to inflammation (Figure 3d). The latter overlap in part with the observations in group 2, e.g. on *TNF and IL−17 signaling*. Of note, immune-related “enriched-up” terms may be confounded by parallel seasonal effects, while the “enriched-down” terms related to a reduction of inflammation are counter-seasonal (Dopico et al., 2015) (their fig. 5).

**Figure 3.**
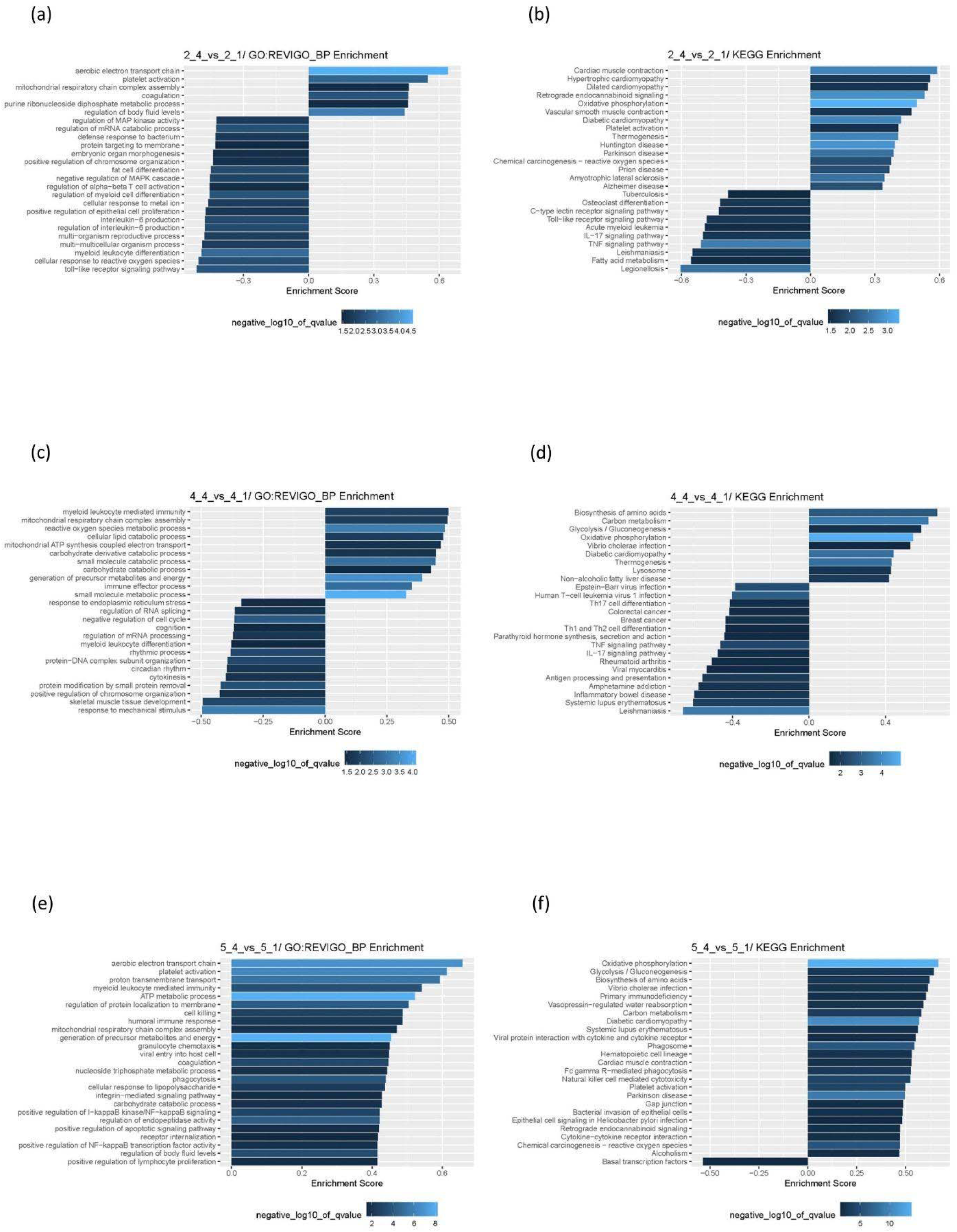
Functional enrichment of the differentially expressed genes, including biological processes (GO:BP, left) and KEGG pathways (right). (a,b) Group 2 longitudinal differences t1 versus t4. (c,d) Group 4 longitudinal differences t1 versus t4. (e,f): Group 5 longitudinal differences t1 versus t4. The size of the bar represents the *Enrichment score* and the color represents the negative log10 of the *q-value* (that is, the adjusted p-value used to control for the false discovery rate), measuring the significance of the enrichment.

Considering our hypothesis that specific seno-therapeutic effects may be obtained due to the SIDs, we were most interested in the longitudinal change of gene expression in the highest-intervention group 5. Since capers in olive oil were consumed in addition to strawberries by group 5, the following enrichments pertain to the whole high-polyphenol diet with three SIDs, which may specifically reflect hormetic responses to the last highest-intervention SID just before the last blood draw. Heatmaps for the GO terms marked by (*) are provided, see Supplementary Data (see listing on page 20). “Enriched-up” in GOBP are terms indicating improvement of mitochondrial (*aerobic electron transport chain* / proton transmembrane transport / ATP metabolic process / mitochondrial respiratory chain complex assembly)*, immune (*myeloid leukocyte mediated immunity* / humoral immune response / granulocyte chemotaxis / positive regulation of lymphocyte proliferation)* and platelet function (*platelet activation\** / *coagulation*), and of apoptosis and associated processes (cell killing / positive regulation of apoptotic signaling pathway* / phagocytosis* / positive regulation of NF-kappaB transcription factor activity) (Figure 3e). All “enriched-down” GOBP terms are below the significance threshold for reporting. For “enriched-up” in KEGG, we again found mitochondrial and metabolism-related pathways (*oxidative phosphorylation*, *glycolysis / gluconeogenesis*), apoptosis-associated processes (*phagosome /* (*Fc gamma R−mediated) phagocytosis* / *natural killer cell mediated cytotoxicity*) and platelet-associated processes (*platelet activation)* (Figure 3f). Of note, *platelet activation* is likely reflecting not an activation, but an increase in the number of cells expressing platelet-specific genes, given the increase in the number of platelets in group 5 (as in all other groups, Suppl. Fig. 1e) and given the dominance of constitutive proteins in the underlying list of genes, including actin beta, integrin subunit alpha 2b, Fc epsilon receptor Ig, myosin light chain, as well as membrane proteins and enzymes. Of interest, the platelet effect was likely counter-seasonal ((Dopico et al., 2015), their fig. 4b), while some immune/inflammation-related terms and also the related term *phagocytosis* may have been confounded by parallel seasonal effects ((Dopico et al., 2015), their fig. 5a; about half of the seasonal phagosomal genes listed in their supplementary fig. 8, that is, CD38, FCGR2A, ITGB2, ITGAM coincide with the genes that we see differentially regulated in the *phagocytosis* KEGG pathway). Finally, KEGG enrichments again highlighted a wide set of (infectious) disease pathways because these feature mitochondrial complexes, tubilin, actin, ubiquitin, histones or immune-related surface marker proteins; the only “enriched-down” KEGG term refers to basal transcription.

#### Harms

No severe side effects were reported by the participants, except for two cases of self-reported strawberry allergy, though without clinical confirmation. Furthermore, 8 participants reported mild intolerance reactions or difficulties consuming the comparatively large amount of extra food on SIDs. (For potential harms of SIDs, specifically regarding potential food-drug interactions, see Box 2).

## Discussion

### A strawberry-based intervention improves lipid profiles and inflammation markers

Here we tested an escalating intervention scheme, over ten weeks, specifically demonstrating the health effects of “seno-intervention days” (SIDs) that feature high amounts of polyphenol-rich food. Our medium-term dietary intervention dose-dependently lowered LDL and CRP in healthy elderly subjects, associated with a specific immunological response. Moreover, effects got stronger as participants continued consumption of the food, demonstrating convincing dose-response relationships. Significant LDL improvement in participants with elevated LDL, as well as mitochondrial improvements and attenuated inflammation (as reflected by the transcriptome) were already triggered by fresh strawberries alone. Adding dried fruit on SIDs, CRP (in participants with a high baseline level) improved. A higher level of dried fruit triggered significant TC improvements (secondary endpoint). Finally, LDL improvements were the strongest, and statistically significant, in the highest intervention group (primary endpoint, *p<0.007*), which also featured putative hormetic effects (specifically in the transcriptome). We can thus claim a health improvement effect in healthy (and well-nourished) elderly people, when considering risk biomarkers as proxies of health. Moreover, for the nutritional intervention tested here, we noted high self-reported compliance and no safety issues, even though the amount of polyphenol-rich food on an SID may be considered quite high by conservative standards, specifically in the highest-intervention group.

The effects of polyphenol-rich diet in general and (straw-)berries in particular are well-known, for both lipid metabolism (specifically LDL and TC) and inflammation (specifically CRP and IL6) (Secci et al., 2021), (Basu, Nguyen, et al., 2014). Many active ingredients in a polyphenol-rich diet, such as quercetin, are considered to mediate these effects, and the host microbiome is involved, since it metabolizes polyphenols (as well as fibers) and also changes in its composition based on diet, giving rise to a complex network of molecular mechanisms (Banerjee & Dhar, 2019). Lists of notable strawberry components are available (Basu, Nguyen, et al., 2014); we singled out quercetin because other polyphenol ingredients of strawberries are less well understood or less frequently reported to be found in notable amounts; furthermore we explicitly identified quercetin in the intervention food (Box 1). In particular, human studies on pelargonidin, the anthocyanin responsible for the red color of strawberries, are scarce; we identified one recent epidemiological study demonstrating neuroprotective effects (Agarwal et al., 2022). Nevertheless, supplementation with anthocyanins has been shown to improve LDL levels in dyslipidemic patients (Qin et al., 2009), while anthocyanin-rich foods are known for some of the effects we found, including improvements of LDL, CRP and TC (Secci et al., 2021). Fisetin is comparatively well-researched, but we could not identify it in the strawberry samples we investigated (Box 1).

Placing our results into context, we focus on human studies, preferably systematic reviews and meta-analyses or summaries thereof, and only for hypotheses about mechanisms we refer to animal or in-vitro studies considering their limited translatability. For (straw-)berries, we will refer to our own recent summary (Secci et al., 2021). For quercetin, we will refer to recent systematic reviews and meta-analyses with a focus on lipid profiles or inflammation, which reported some specific yet conflicting evidence for effects. Overall, quercetin effects on lipid profiles were found to be limited to triglycerides (Sahebkar, 2017), to HDL and triglycerides following interventions of more than 8 weeks (Huang, Liao, Dong, & Pu, 2020) and to LDL and TC in case of metabolic syndrome and related disorders (Tabrizi et al., 2020). Anti-inflammatory effects of quercetin in terms of CRP reductions were found (Mohammadi-Sartang, Mazloom, Sherafatmanesh, Ghorbani, & Firoozi, 2017), sometimes limited to subgroups such as subjects with diagnosed diseases or metabolic syndrome (Tabrizi et al., 2020), also for IL6 (but not for TNFα) (Ou, Zheng, Zhao, & Lin, 2020).

In our study, the positive cholesterol-related effects were stronger for TC than for LDL in all intervention groups, with the effects being visible in group 2 (fresh strawberries only) and increasing monotonously in size with each escalation step for both markers (Table 2), up to −0.148 mmol/L in case of LDL and up to −0.25 mmol/L in case of TC. Such a clear dose-response relationship is not always found; most recently it was reported that the lower dose, but not the higher dose of strawberry powder was effective in reducing LDL, for unclear reasons (Richter et al., 2022). HDL and triglycerides showed no intervention-related pattern in our study; however HDL improved overall.

In detail, past meta-studies reported cholesterol effects for (straw-)berries (Secci et al., 2021), and average LDL reductions were between −0.21 mmol/L (Huang, Chen, Liao, Zhu, & Xue, 2016) and −0.23 mmol/L (−8.86 mg/dL) (Shah & Shah, 2018), while average TC reductions were −0.17 mmol/L (−6.49 mg/dL) (Hadi et al., 2019). For quercetin in particular, one meta-analysis found LDL and TC reductions in people with metabolic syndrome or related disorders (Tabrizi et al., 2020). Thus, the reduction of LDL (up to –0.148 mmol/L) and of TC (up to –0.25 mmol/L) in our study is comparable to what was reported in the literature. Mechanistically, cholesterol effects by (straw-)berries and a polyphenol-rich diet in general, and by quercetin in particular may be explained by the inhibition of cholesterol uptake through competition and by the direct influence on membrane properties and cellular signaling cascades (Jia et al., 2019). Interestingly, the decreases in TC in our study were even more pronounced than in LDL cholesterol, which could (together with positive effects on HDL) point to a decrease of atherogenic small dense LDL and/or other atherogenic particles. The downregulation of PCSK9 and the upregulation of ABCA1 shown in mice after quercetin treatment are other possible explanations for lower LDL (and increasing HDL) (Jia et al., 2019). Also, phytosterols from strawberries have been shown to lower LDL and LDL-like particles by competing with cholesterol absorption (Basu, Nguyen, et al., 2014). Cardiovascular diseases are still one of the main causes of death in industrialized countries, with disorders of lipid metabolism playing a central role. Lowering LDL with statins has side effects and therefore additional dietary LDL lowering both in primary and secondary prevention is important to reduce required statin doses.

Inflammation-related effects were visible in our study, although these are more dynamic. More specifically, the interventions (in particular the SIDs in groups 3-5) may feature acute versus chronic effects in time, in a hormetic fashion. Yet, in subjects with initially elevated CRP values we observed a strong approximately 50% decrease in CRP (by up to around –0.7 mg/L) already after the first two weeks of intervention, but only in the SID groups 3-5. For IL6 we observed a decrease in all groups overshadowed by an intervention effect, in group 5 only, where the decrease was significantly less than in all other groups, including group 1. We thus postulate that in all high-intervention groups, a chronic anti-inflammatory effect is present, while in group 5, an acute immune reaction with a relative increase in inflammatory markers co-occurs one day after the SID, possibly triggered by apoptotic/senolytic processes in blood cells. This scenario is motivated and supported by the gene expression data, see below. Overall, the anti-inflammatory pattern may be further influenced by seasonal effects: these would have been in a pro-inflammatory direction (Dopico et al., 2015) for the duration of our study, making an even stronger anti-inflammatory effect of the intervention necessary to yield a visible anti-inflammatory effect.

In detail, in the past, anti-inflammatory effects by (straw-)berry interventions (Secci et al., 2021) were reported to average −0.33 mg/L (Fallah, Sarmast, Fatehi, & Jafari, 2020), −0.47 mg/L (Hadi et al., 2019), and −0.63 mg/L (Gao, Qin, Arafa, Eshak, & Dong, 2020) for CRP, and −0.41 pg/ml for IL6 (Fallah, Sarmast, Fatehi, et al., 2020). Reductions by quercetin interventions averaged −0.33 mg/L (Mohammadi-Sartang et al., 2017) and –0.11 mg/L for CRP (Ou et al., 2020), and –0.58 pg/mL for IL6 (Ou et al., 2020). Thus, the anti-inflammatory effects we report for CRP (up to –0.7 mg/L) are comparable to the literature. We also witnessed comparable reductions of IL6 between −0.5 to −1 pg/mL, yet we were surprised that these were also found in the control group. We have no explanation for this finding, although we can explain the only moderate reduction of IL6 in the highest-intervention group, see below. Mechanistically, a plethora of anti-inflammatory effects are attributed to (straw-) berries / polyphenols in general and quercetin in particular, and they center around an inflammatory cytokine axis featuring TNFα, IL6 and CRP (Secci et al., 2021), (Ou et al., 2020),(Serino & Salazar, 2018). CRP is an important acute-phase protein of hepatic origin that binds to lysophosphatidylcholine expressed on the surface of certain types of bacteria or dying cells to activate the complement system and to eliminate these bacteria as well as dead cells. The role of CRP and its mediators is difficult to pin down and multifunctional. For example, in combination with complement components, CRP can promote the noninflammatory clearance of apoptotic cells (Gershov, Kim, Brot, & Elkon, 2000); CRP-induced apoptosis was also observed (Blaschke et al., 2004). Of note, CRP is known to be inhibited by quercetin in vitro, (Garcia-Mediavilla et al., 2007). Moreover, strawberry extract and pelargonidin-3-O-glucoside were shown to downregulate TNFα and IL6 in a mouse model of inflammation, and in vitro, pelargonidin-3-O-glucoside showed similar effects and also inhibited NFκB and MAPK signaling (Duarte et al., 2018). Testing strawberry extract effects ourselves, we stimulated cultured hepatocytes with IL1β/IL6, triggering a manyfold increase of the CRP mRNA level compared to untreated cells, and a water-soluble strawberry extract indeed impeded CRP induction in a dose-dependent manner, see Box 3. Furthermore, in the gene expression data (see below), anti-inflammatory changes were found, with a superimposed immune-inflammatory hormetic effect in the highest-intervention group. As an aside, and supported by the gene expression data, we found an increase of platelet number within the healthy physiological range for all groups (Suppl. Fig. 1e), despite the seasonally expected decrease (Dopico et al., 2015); this increase is moderate and far within the U-shaped reference range positively predictive for health (Tsai, Chen, Lin, Huang, & Tarng, 2015) and a variety of *anti-inflammatory* effects of platelets were described in addition to pro-inflammatory ones (Ludwig, Hilger, Zarbock, & Rossaint, 2022).

There are numerous interactions between the negative (atherogenic) changes in lipid metabolism and inflammatory processes that reinforce each other. Thus, the changes shown here are likely physiologically more positive than any changes in the individual factors per se (Johnson & Stolzing, 2019), (Walter, 2009), (Ademowo, Dias, Burton, & Griffiths, 2017). CRP colocalizes with and binds to LDL phospholipids, particularly when the structure of CRP is subtly altered by oxidation. Moreover, CRP is a ligand of lectin-like oxidized LDL receptor-1 (LOX-1) (Fujita et al., 2009) and increases LOX-1 expression in human aortic endothelial cells (Li, Roumeliotis, Sawamura, & Renier, 2004) and thus enhances the uptake of oxidized LDL. Such interplay with LOX-1 and atherogenic LDL may enhance endothelial dysfunction and additionally promote atherogenesis (Stancel et al., 2016). Due to these synergistic effects of LDL and CRP, moderately elevated CRP values belong to the so-called “emerging” risk factors, which for values above 2 mg/L indicate a need for treatment, even in case of only moderately elevated LDL (Ridker et al., 2008), (Choudhry, Patrick, Glynn, & Avorn, 2011). In many epidemiological studies, LDL and CRP have been shown to have a synergistic effect for both myocardial infarction (Mo et al., 2020), (Ridker et al., 2005) and stroke (Kitagawa et al., 2019).

### A strawberry-based intervention improves mitochondrial, immune and platelet function, and may clear senescent blood cells

The GOBP and KEGG findings for groups 2 and 4 (group 3 was not investigated) yielded many processes and pathways demonstrating improvement of mitochondrial, immunity and platelet function, as well as suppression of inflammation; these observations match the hypothesis that a strawberry-based polyphenol-rich intervention may not just attenuate inflammation as described above, but may also improve mitochondrial metabolism (Vasquez-Reyes et al., 2021), (Chodari et al., 2021) and immunity (Sangwan, Farag, & Modolo, 2022), (Pahwa & Sharan, 2022), (Basak & Gokhale, 2022). For the highest-intervention group 5, gene expression-based enrichments suggest a more complex scenario, as follows. 1) Mitochondrial energy metabolism is enhanced in many ways, potentially contributing to better immunity and hormetic inflammation. Preclinical and a few clinical studies specifically support mitochondrial improvements by polyphenols (Vasquez-Reyes et al., 2021), (Chodari et al., 2021). 2) Immunity is enhanced, including *myeloid leukocyte mediated immunity, phagocytosis,* and *platelet activation*. We also found an enrichment of *cell killing* and *positive regulation of apoptotic signaling pathway,* associated with *positive regulation of NFkB activity*. This may be an indication of senolysis of senescent blood cells, with a corresponding hormetic activation of some inflammation and immunity processes. Accordingly, IL6 *protein* expression went down the least in group 5, compared to group 1. Furthermore, in vitro, quercetin alone (as well as oleuropein from olive oil) displays anti-senescence activity (Maria & Ingrid, 2017),(Varela-Eirin et al., 2020), while also triggering apoptotic and anti-proliferative activity in tumor cells (which are, however, not usually senescent) in-vitro and in some animal models (Ghafouri-Fard, Shoorei, Khanbabapour Sasi, Taheri, & Ayatollahi, 2021). 4) We also find an enrichment for the term *phagocytosis*, which may be an event downstream of apoptosis. Alternatively, though, *phagocytosis* and related processes may just be downstream of the immune activation we observe in all high-intervention groups. Correspondingly, in the context of bovine mastitis, quercetin was shown to enhance *phagocytosis and bacterial killing* (Disbanchong et al., 2021) and it promotes the expression of genes involved in phagocytosis in bovine neutrophils (Srikok, Nambut, Wongsawan, & Shuammitri, 2017). Overall, the gene expression data align with the inflammation marker data, adding mitochondrial and immune aspects, and they suggest apoptosis and related processes triggered in the highest intervention group, possibly due to the senolysis of senescent blood cells.

In terms of gene expression data, as the closest match to any meta-analysis, an integrative analysis of five studies reported positive effects of polyphenols on cardiometabolic health in humans (Ruskovska et al., 2022), implicating processes such as cell adhesion and mobility, immune system, metabolism, or cell signaling. More specifically, *TNF and toll−like receptor signaling* and *oxidative phosphorylation* were found, as we did for the high-intervention groups 2 and 4, and *natural killer cell mediated cytotoxicity* and the *phagosome* as we did for the highest-intervention group 5. A few studies have investigated the effects of an intervention with quercetin on human gene expression, while (straw)berry studies are lacking. In particular, Boomgaarden et al. (Boomgaarden et al., 2010) showed that daily supplementation with quercetin in healthy individuals led to changes in the gene expression profiles of human monocytes; 62 subjects were enrolled in two studies. Overall, quercetin triggered small yet significant changes in the expression of genes related to immune function, nucleic acid metabolism, and cell death, matching some of our findings.

### Limitations and Strengths: Nutritional trial interpretation and potential sources of bias

Nutritional geroprotective interventions are attractive because of the outstanding safety attributed to changes in diet. A long history of dietary changes due to human and environmental evolution and adaptation as well as more recent epidemiological data confirm that effects, positive as well as negative ones, are usually small. We set out to test how modest as well as more radical changes of diet may affect health when aiming for a specific polyphenol-rich diet, adding strawberries and capers in olive oil to the standard diet of healthy elderly people. We note that any diet intervention is poorly defined because the exact composition of food in terms of ingredients can be quite variable. We checked food samples for two polyphenols, identifying quercetin but not fisetin (Box 1). We expect that a plethora of active ingredients contributed to the effects we are reporting, potentially in synergistic ways (van Breda & de Kok, 2018).

Our selected study population was already featuring a high level of health at the start of the study, as demonstrated by their *Phenotypic Age*, which was already ∼6-8 years younger than reference at baseline (Table 1, Fig. 2e). Nevertheless, we observed multiple health improvements in all groups (*Phenotypic Age,* EQs, HDL, IL6, sTNFr1, GDF-15, Chair-Rise-Test); just participating in the study (inclusion bias) may have triggered these positive effects. Moreover, placebo effects need to be considered in nutritional studies. No placebo was employed here, as fresh strawberries cannot be “faked”. In fact, the instructions to participants explicitly stated that the intervention food was chosen based on well-known knowledge about its health-promoting effects. We thus acknowledge that all effects reported are a mix of true intervention effects and placebo (and other confounder) effects. Another source of bias in dietary intervention studies are replacement effects, e.g. the high strawberry intake may have implied a lower intake of other fruit (and vegetables), but it may as well have implied a lower intake of less healthy foods, and we did not attempt to find out. After ten weeks, the point estimate for body weight change was –0.10 kg (*p=0.433*), with no group-specific differences, rendering it unlikely that effects observed were due to caloric restriction. The analysis of potential effect modifications was generally limited by statistical power, since it involves 3-fold interaction terms in the LMM. Nevertheless, sex may have modified the effect of the intervention on LDL (*p=0.011* for the three-fold interaction group:visit:sex), and the difference in LDL change to baseline between women and men was highest in group 5 (up to –0.472 mmol/L at visit 3), reflecting a larger decrease in women. We found no other specific effects of age group, sex, smoking or marital status.

While some effects may at least in part be due to bias, it is also possible that effects were missed, for example by an insufficient coverage of effects by the proxy biomarkers or sampling timepoints. Detecting the effects of the SIDs is particularly challenging; we sampled blood one day after the SID intervention and we found evidence that hormetic inflammatory responses were still dominating by then. The SID response is expected to be a mix of responses, and the comparatively high values of IL6 in the highest-intervention group hint at a temporary inflammatory peak, potentially triggered by the apoptosis of cells and downstream consequences, such as phagocytosis (see, e.g. (Barnes, 2021)), and suggesting that polyphenols (and, specifically, the large amount of quercetin) of the SID may have triggered the apoptosis of senescent blood cells. Since the amount of senescent cells is usually limited to a small percentage of all cells, the resulting elimination of cells is not expected to be visible in standard cell count data. The strengths of our study lie in the large sample size, exceeding previous reports for strawberries, in the dose-escalation design, and in the intervention foods and biomarker measurements selected specifically based on our hypotheses regarding the role of lipids, inflammation and apoptosis/senescence for (cardiovascular) health.

### Conclusion

We found that a relatively short-term but intense diet with polyphenols can significantly lower LDL and trigger an overall more favorable cardiovascular profile, and potentially a senolytic response. To extend healthy life expectancy, good nutrition, exercise and socio-psychological interventions tend to feature few side effects as well as easy availability under most circumstances. Optimizing these interventions towards a maximum of synergy, ideally based on easily affordable biomarkers, and thus tailoring them to specific subpopulations or even specific individuals requires observational and interventional studies. Our study contributes some insights towards designing and selecting nutritional interventions for maximum effect, and encourages future work to define high-effect nutritional interventions.

#### Box 1.

##### Quercetin (and fisetin) contents in strawberries, capers and controls

We employed mass spectrometry to measure the amounts of quercetin and fisetin in strawberries of two of the varieties consumed by the participants, that is, *Malwina* and *Florentina* (the latter measured twice in August 2021, two weeks apart), see Supplementary Text (see listing on page 20). We found approximately 0.35-1.04 µg/g of quercetin, but β-glucuronidase pretreatment resulted in amounts of up to 40 times higher, of approximately 3.91-14.07 µg/g. Since β-glucuronidase is expressed by human cells (Gervasi et al., 2022), (Barreca et al., 2021) we assume that these higher amounts of quercetin are bioavailable in principle. In capers, we found 46.1 µg/g before, and 108.4 µg/g after β-glucuronidase pretreatment; we also found quercetin in lovage samples (positive control). We could not measure any of the quercetin from the capers that may have been dissolved in the olive oil during storage. We failed to detect fisetin in strawberries, yet we found it in smoketree samples (positive control). The amount of quercetin (canonically measured without β-glucuronidase pre-treatment) reported in the literature varies widely, for strawberries as for other berries, and also for capers (see, e.g., (Basu, Nguyen, et al., 2014), (Azzini et al., 2010)); measurement variability is certainly an issue here. Natural variation is an issue as well, however.

We expect that polyphenol amounts in general, and quercetin amounts in particular, thus varied widely in the food consumed by our participants, depending, e.g., on the exact variety being consumed, on the day the fruits were harvested, and for how long and how the fruits were stored before eating. 100g freeze-dried strawberries were derived from approx. 1.0-1.2 kg of fresh fruit; we did not measure quercetin in the dried fruit, but strawberry powder based on freeze-drying was used in past trials and is known to still contain high polyphenol contents (Basu, Nguyen, et al., 2014). Moreover, we only measured the quercetin aglycone and glucuronidated quercetin, while there are a lot more quercetin derivatives found in food (Dabeek & Marra, 2019). Given all the unknowns mentioned, we refrain from any attempts to estimate the amount of quercetin consumed by the various groups in our study; total polyphenol content would be even more difficult to estimate.

Finally, in the plasma of participants of the pilot trial, we could not detect quercetin (similar to (Azzini et al., 2010)), nor fisetin, see Supplementary Text (see listing on page 20). Given the half-life of quercetin, we expect that the quercetin available in the food was mostly modified or degraded by metabolism by the time the blood samples were taken (Dabeek & Marra, 2019), (Robbins et al., 2021).

#### Box 2

##### Side effects and toxicity

Apart from potential allergies to components of the dietary intervention that we investigated (see main text), polyphenol toxicity and the comparatively high amount of sugar in strawberries were considered as potentially detrimental effects of the intervention. For both, it may theoretically be the case that the natural safety-net of the digestive system (loss of appetite, aversion to the food in question, vomiting or diarrhea) do not perform sufficiently well in certain people. In particular, polyphenol toxicity is occasionally mentioned in the literature, but mostly based on in vitro or animal work. A comprehensive review from 2012 noted concerns regarding iron absorption, pro-oxidant effects and competition for enzymes that may trigger drug interaction effects (Hooper & Frazier, 2012); green tea polyphenols were specifically mentioned as lookalikes of folic acid, and soy polyphenols of hormones. However, iron absorption is supposedly a problem if polyphenols are not consumed together with vitamin C, which is not the case here. Enzyme inhibition of, e.g. the cytochrome P450 enzymes also needed for drug metabolism, was reported for grapefruit and cranberries, but was not reported yet for strawberries, capers or olive oil. However, it was reported that quercetin and digoxin may compete for the P-gp (P-glycoprotein) transporter, causing lethal events in pigs (Wang, Chao, Hsiu, Wen, & Hou, 2004). Also, cyclosporin, vinblastine, vincristine and nephrotoxic drugs are contraindicated, as is warfarin (Lopes et al., 2021), (Lopez-Yerena et al., 2020). Of note, use of these drugs was incompatible with inclusion into our study.

Quercetin is approved by the US Food and Drug Administration as GRAS (Generally Recognized As Safe) for use as an ingredient at levels up to 500 mg per serving (Barreca et al., 2021) (Pawar et al., 2022). In fact, with no reported side-effects, doses of 500 mg or 1000 mg were consumed daily over 12 weeks by ∼600 adults (Knab et al., 2011), 1000 mg of quercetin daily were consumed over 6 weeks by ∼20 healthy adults (athletes) (McAnulty et al., 2008), and doses from 250 mg to 5000 mg of quercetin daily over 4 weeks, by 30 chronic hepatitis C patients (Lu et al., 2016). Moreover, quercetin (up to 600 mg daily) was suggested by some research groups to be a supplement of choice for COVID-19 and associated comorbid conditions (Pawar et al., 2022). Specific toxicity incidents related to anthocyanins are lacking, probably due to their very low bioavailability (Wallace & Giusti, 2015).

Further, pro-oxidant effects may be triggered by polyphenols, and for these we argue that such effects, if any, should be considered to be welcome because these are then a hormetic cause for improvements. We thus hypothesize, as was done for exercise, caloric restriction and many other healthy interventions (Demirovic & Rattan, 2013), that a high-polyphenol intervention may actually trigger a minor yet notable challenge to the participants, and it is the hormetic repair response which results in health improvements. More specifically, the high dose in particular in group 5 may have caused inflammation or apoptosis of e.g. senescent cells. In fact, in self-experiments (N=4) in early 2020, we did close-meshed blood draws (every 4 hours and next morning) and found some cytokine levels to increase in the 8 hours after the intervention, returning to baseline level next day, but these observations were not significant, and were hard to interpret given that cytokines are also subject to circadian rhythms that are not well-studied. A more recent review reiterated some of these aspects (Cory, Passarelli, Szeto, Tamez, & Mattei, 2018) and suggested to favor diet over supplements. Finally, (Hernández & Gil, 2021) [Chapter 69 therein] do not add further concerns, but stress the need to go beyond in vitro and animal experiments, noting the lack of clinical trials. These authors also noted that “No beneficial effects have been observed for antioxidant supplementation in well-nourished people with a balanced diet. However, a high intake of polyphenols, even from dietary sources, can result in toxic effects (Das et al., 2012)”, yet the review by (Das, Bhaumik, Raychaudhuri, & Chakraborty, 2012) is based on preclinical data, and our results suggest that healthy people can profit, at least in terms of reducing risk factors.

Regarding the sugar content of strawberries, we note first of all that overt diabetes (requiring prescription of insulin) was an exclusion criterion per se. Furthermore, we considered early on that strawberries have been tested successfully multiple times as the basis of *treating* people with metabolic syndrome and obesity (Basu, Betts, et al., 2014), and quercetin as well as anthocyanins were reported to be useful for diabetes management (Cory et al., 2018), (Barreca et al., 2021); presumably the polyphenol or fiber content of strawberries more than outweigh any problems due to the sugar content of the fruit. Regarding the capers in olive oil, we noticed the high content of salt in the food we provided to the participants, and instructed them to reduce the consumption of salty food on the days on which capers were consumed. Further, elevated values of triglycerides may be the result of a higher consumption of olive oil (Jabbarzadeh-Ganjeh, Jayedi, & Shab-Bidar, 2022). Finally, the hypothesis of synergistic positive effects of our food intervention may refer to synergistic toxic effects as well, and the combination of strawberries, capers and olive oil may not be considered a time-tested combination even though it matches the mediterranean diet scheme (Esposito et al., 2022). We thus successfully tested the combination before the trial, in *N=1* and *N=4* self experiments and in the pilot trial of 19 participants in Summer 2020, further minimizing the chance that our food combination may be synergistically toxic and yet no knowledge about toxicity exists because the intervention was not done frequently in the past.

#### Box 3.

##### In-vitro testing of strawberry extract in hepatocytes expressing CRP

Stimulating cultured hepatocytes with the recombinant interleukins 1β and 6 (IL1β/IL6) resulted in a 4.2-fold increase of *CRP* mRNA level compared to untreated cells (see Figure). This CRP induction was significantly impeded by the presence of a water-soluble strawberry extract in a dose-dependent manner. The highest concentration tested (5 mg/ml) also showed the highest inhibitory activity (90 % inhibition of IL1b/IL6-dependent *CRP* mRNA induction). The lowest concentration of the strawberry extract (5 µg/ml) still exhibited a non-significant inhibitory activity of 33 % (see Figure); see Supplementary Text (see listing on page 20) for Methods.

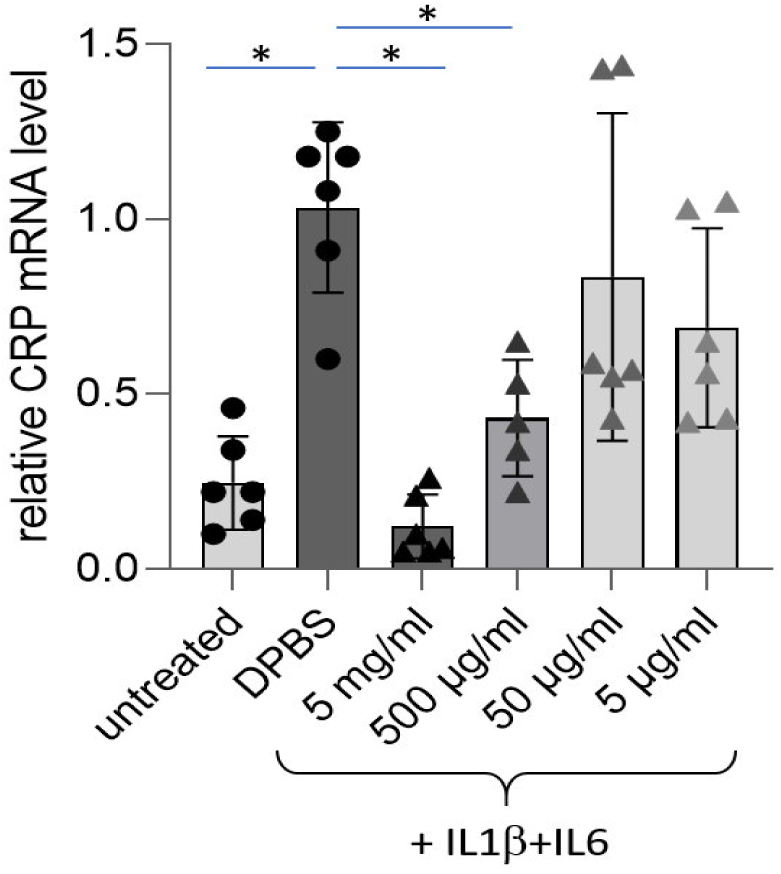

Figure. Interleukin-dependent CRP induction in human cultured hepatocytes is reduced in presence of a water-soluble strawberry extract. The *CRP* mRNA level was assessed with quantitative RT-PCR and related to the housekeeping gene expression of *GAPDH*. The mean of the solvent control group (DPBS) was set to be one. Cell culture experiments were performed in duplicates, for three times. Individual data, means and standard error of the mean are displayed.

## Supporting information

Supplementary Methods Results and Discussion

## Data Availability

Aggregated data that do not require managed access are provided in the Supplementary Files (see listing on page 20). Other data are available from the authors upon request.

## Acknowledgements

The study was funded in part by Karls Erdbeerhof (Karl’s strawberry farm). We also thank the Interdisziplinäre Fakultät of the University of Rostock (Altern des Individuums und der Gesellschaft) for intramural financial support. We thank Siemens Healthcare for donating reagents. We thank EFRE (European Fund for Regional Development) [GHS-15-0019, 2016-2021] and the *Core facility for Cell sorting and Analysis* for infrastructural support. The mass spectrometric analyses of quercetin and fisetin were performed with a triple quadrupole mass spectrometer LCMS-8050 co-funded by the Deutsche Forschungsgemeinschaft (DFG, INST 264/169-1 FUGG). The analysis of micronutrients was funded by the BMBF (FKZ 031B0731E) as part of the joint project “f4f—food for future”. G.F., A.K. and D.P. are further supported by the BMBF (FKZ 01ZX1903A and 1DS19049). We thank all participants, all expert advisors consulted before the study, and all helpful people at Karls. We also thank the local teams at the IBIMA and ILAB (A. Brauer, F. Thiesen, S. Schenk, R. Krause, E. Kauffold, L. Werner, A. Werderitsch, A.-L. Roßmann, R. Müller, Z. Setayeshmehr, C. Raßmus, A. Beier, T. Freitag). The funders had no role in study design, data collection, data analysis, data interpretation, or writing of the report.

## Author contributions

A.H. and M.W. implemented the clinical study. R.Se. analyzed the gene expression data. H.R. contributed the biostatistics analysis of the clinical data. J.M. and K.J. contributed to implementing the study. A.H., E.S.-T., I.B., D.P. and A.K. contributed to analyzing and interpreting the data. R.Schw. and B.H. contributed the mass spectrometry data, analysis and interpretation. D.W. and T.G. contributed the micronutrient and MDA data, analysis and interpretation. P.H. and G.R. contributed the in-vitro data, analysis and interpretation. G.F., M.W. and H.R. conceived the study, analyzed and interpreted the data and wrote the manuscript with contributions of the other authors.

## Supplementary Files

**In the following, all Supplementary Files, Data, Figures and Texts are listed and described**

### A) Supplementary Files (Ethics and administrative matters, available at DRKS, the German Clinical Trials Registry, or from the corresponding authors)

1. **Ethics Application, Study Protocol** (03/2021, in German, <u>available at DRKS</u>); the Ethics Application, Study Protocol, tracked changes (in German, based on the ErdBEHR1 pilot study with Ethics Approval in 2020, is <u>also available at DRKS</u>)
2. **Ethics Approval** (04/2021, in German, available upon request from the corresponding authors)
3. **Supplementary Documents** (in German, available upon request from the corresponding authors)
  a. Recruitment questionnaire
  b. Info sheet (Informationsblatt) ErdBEHR-2-Studie
  c. Clinical data questionnaire (Klinischer Fragebogen ErdBEHR-Studie)
  d. 2nd Clinical data questionnaire (Zweiter Klinischer Fragebogen)
  e. Detailed personalized consumption plan, sample (Verzehrplan Gruppe 5)
4. **<u>Recipes given to the participants (google drive)</u>** (in German, also available upon request from the corresponding authors)

### B) Supplementary Data

1. **PCA** The “PCA” <u>folder</u> (google drive) presents the results of the Principal Component Analysis (PCA) of the gene expression samples, focusing on the top 500 most variable genes.
2. **DEG** The “DEG” <u>folder</u> (google drive) contains the results of the differential gene expression (DEG) analysis, which compares the gene expression profiles between different groups. The analysis results are aggregated in the file differentialtable.csv. In addition, there are other files that contain the specific DEGs for each comparison, and their names indicate the comparison groups. These comparisons included all groups (1,2,4 and 5) at timepoint 4 vs timepoint 1, as well as group 5 vs group 1 at timepoint 1 and at timepoint 4.
3. **Enrichment_GSEA** The “enrichment_GSEA” <u>folder</u> (google drive) contains the results of the Gene Set Enrichment Analysis (GSEA), based on the DEG data. The results are organized into group-specific subfolders. Each subfolder includes bar plots for the REVIGO and KEGG results, as well as tables that provide the GO, KEGG, and REVIGO results used to generate the plots.
4. **Heatmaps** The “heatmaps” <u>folder</u> (google drive) contains heatmaps created from the gene expression data. The heatmaps show the gene expression values of a specific subset of the genes across the samples, as follows. There are heatmaps of the genes belonging to a selected GO term (see main text; myeloid leukocyte mediated immunity, platelet activation, phagocytosis, aerobic electron transport chain, positive regulation of apoptotic signaling) which is specified in the title of each heatmap. Note that no measurements were done for group 3. Transcript abundance is visualized as the binary logarithm of the read count.

### C) Supplementary Figures

**Please see the end of the Main Manuscript.**

### D) Supplementary Methods, Results and Discussion

1. **Presentation of the group 1 gene expression data.**
2. **Box 1 Supplementary Results, Discussion and Methods.**
3. **Box 3 Supplementary Methods.**

**Please see the Supplementary Methods, Results and Discussion in a separate file.**

---

**Suppl. Fig. 1.**
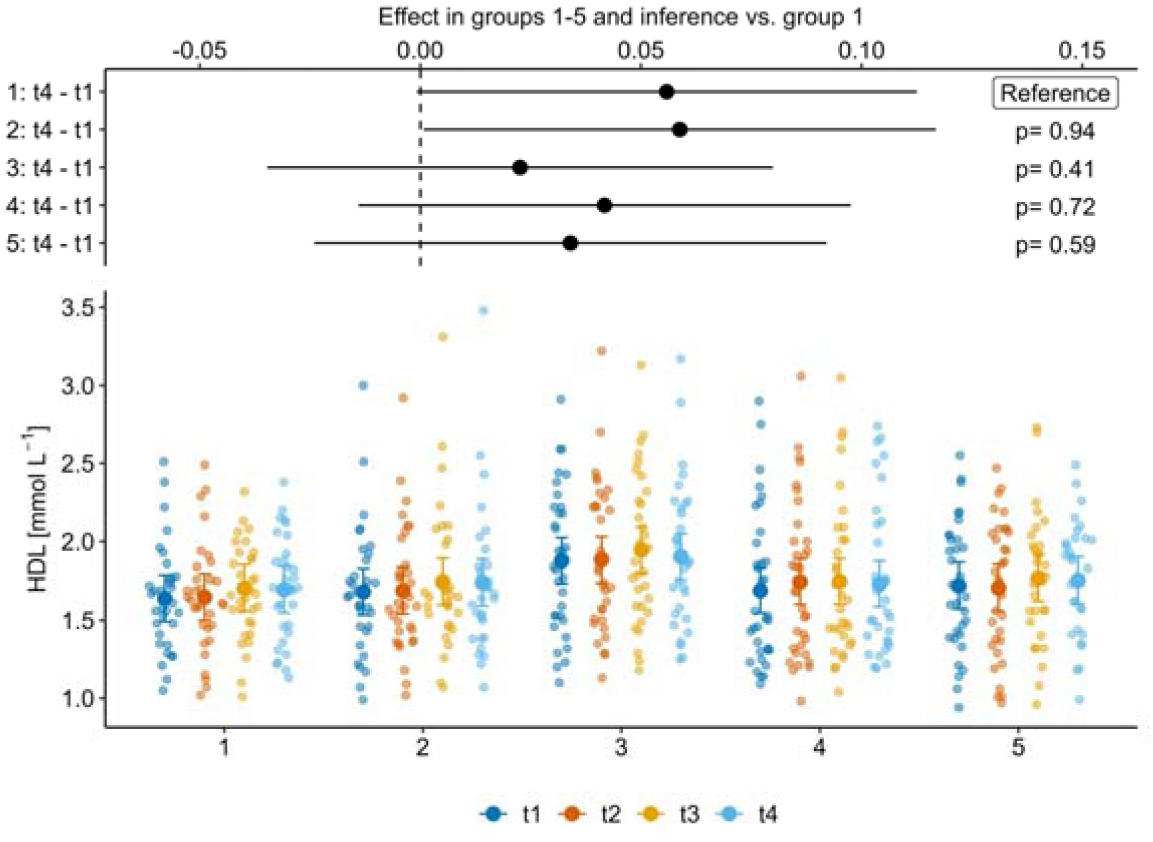
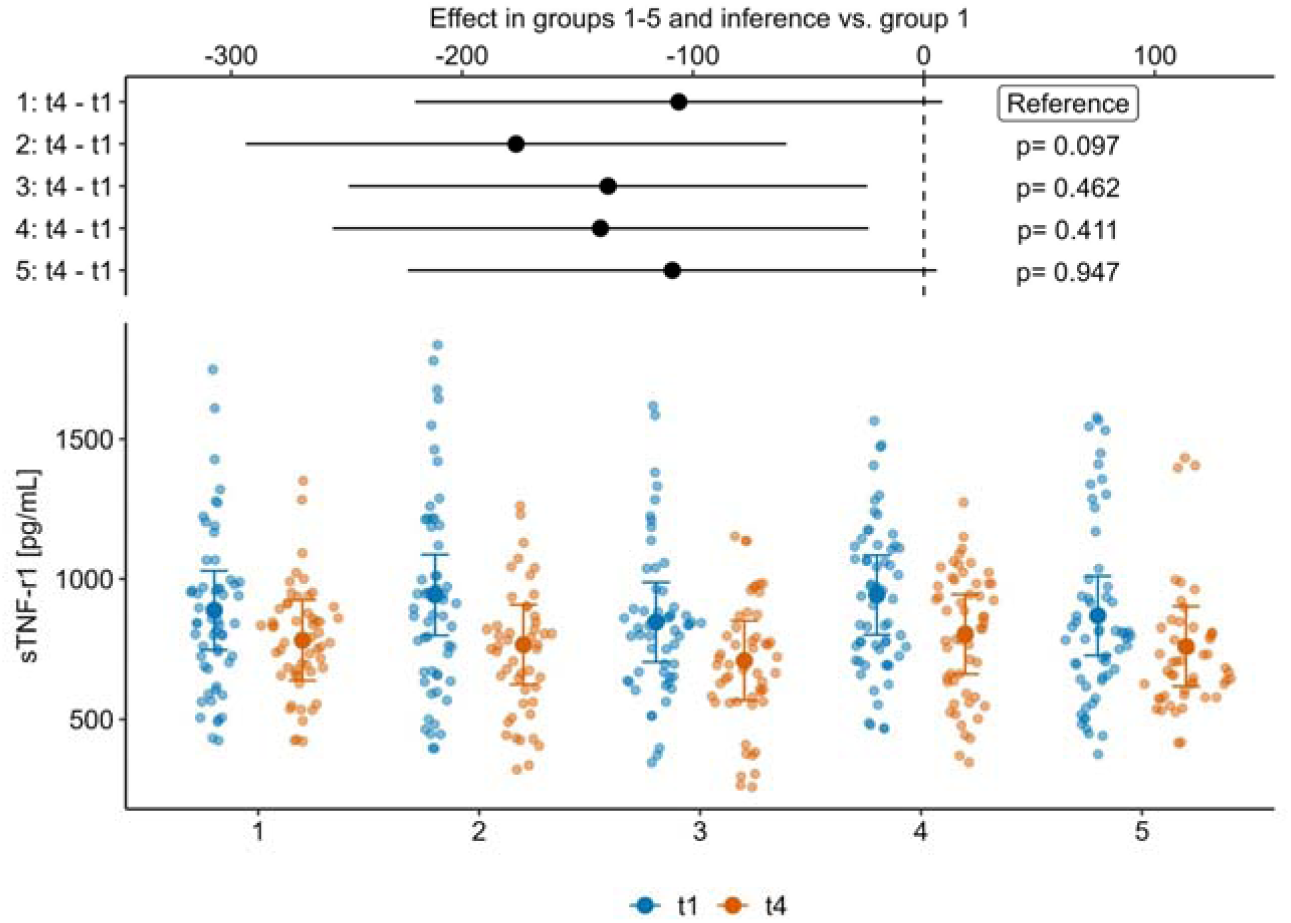
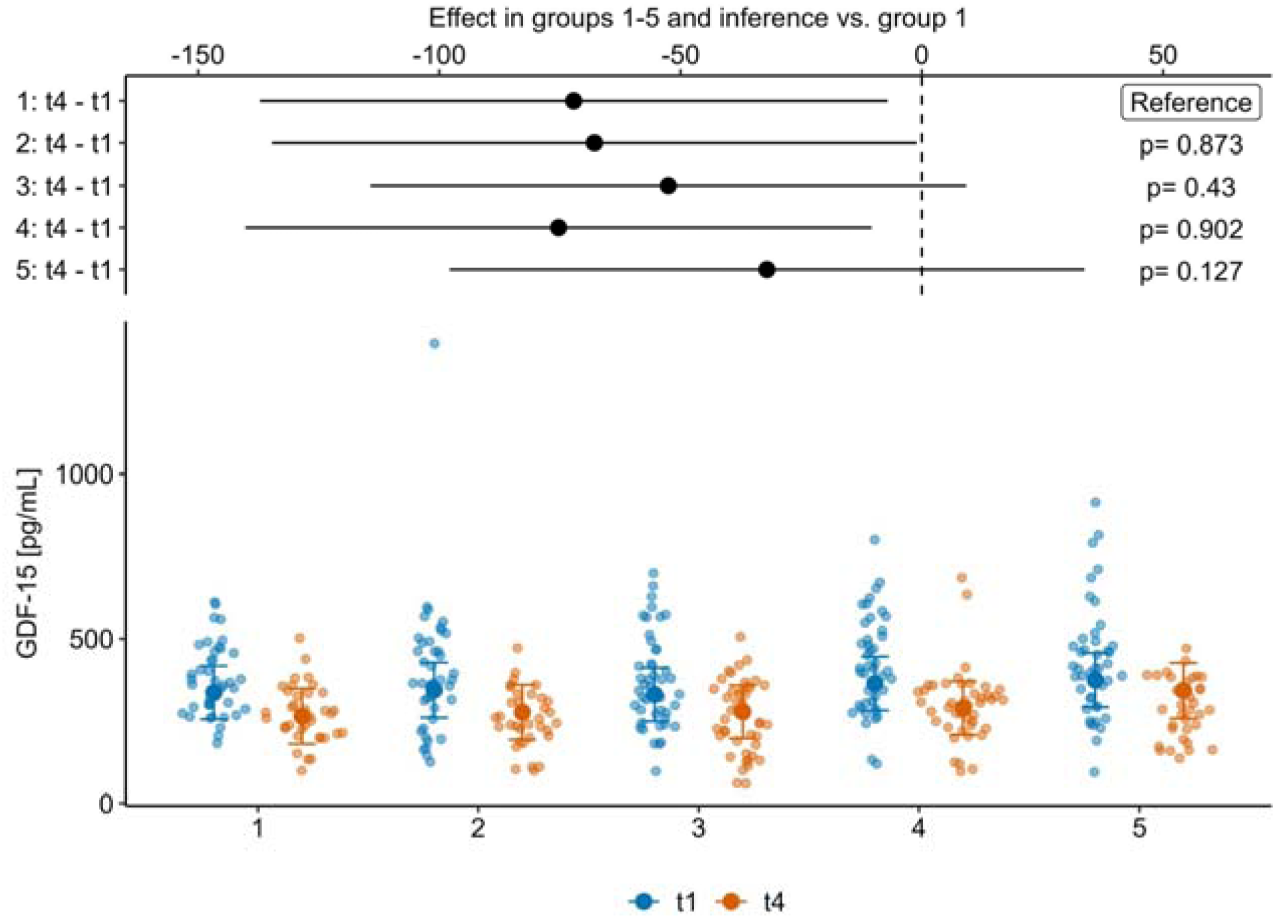
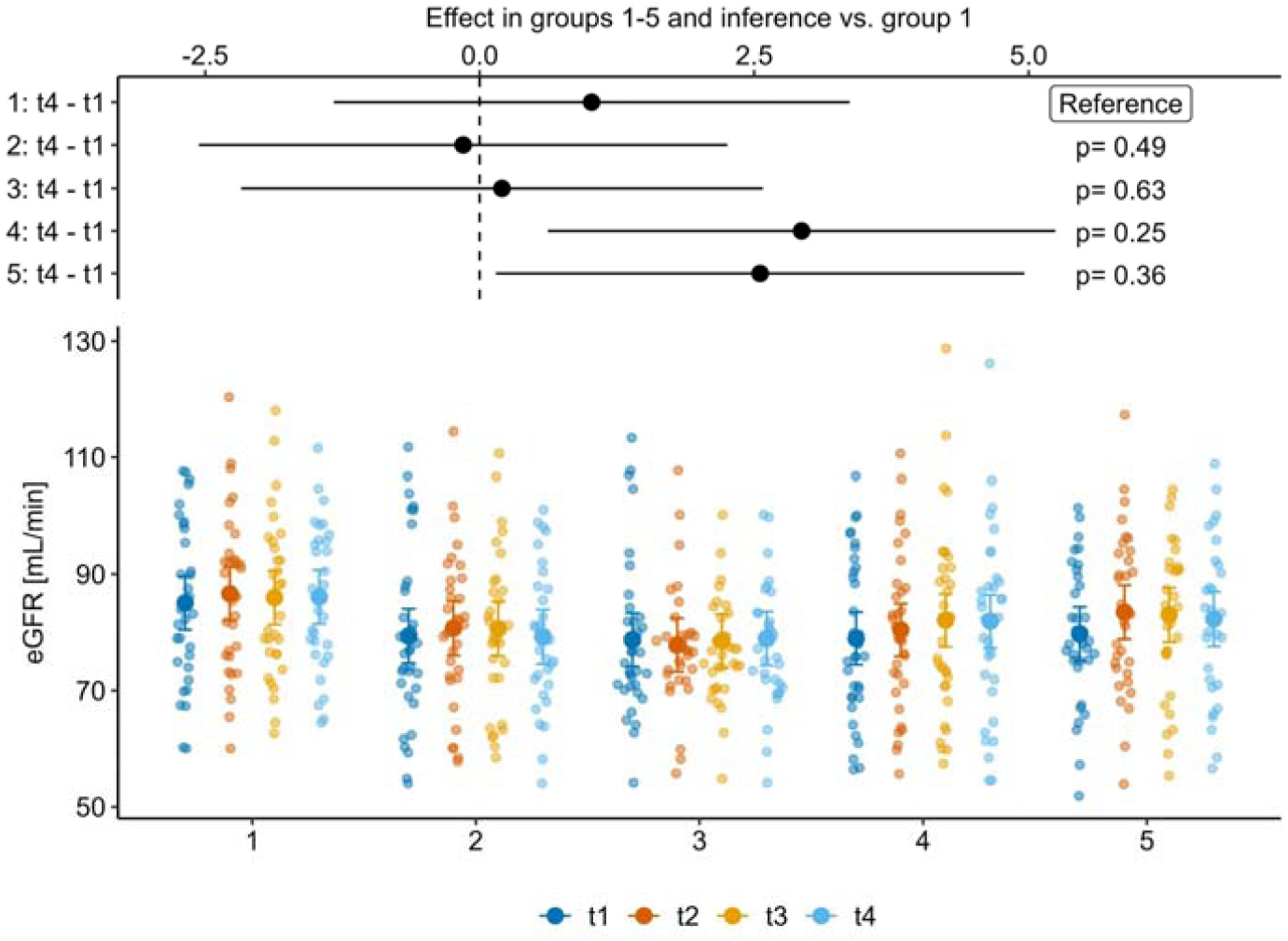
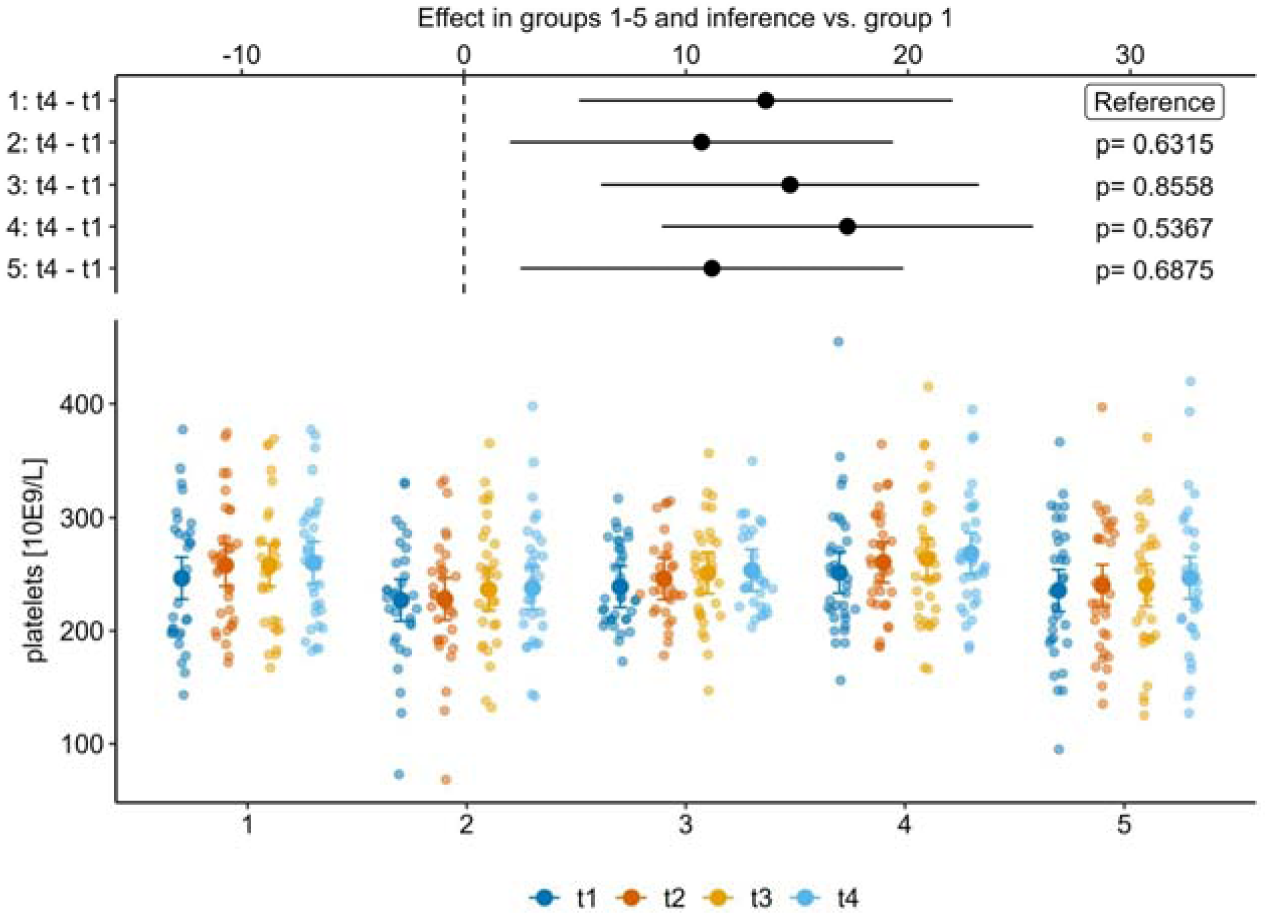
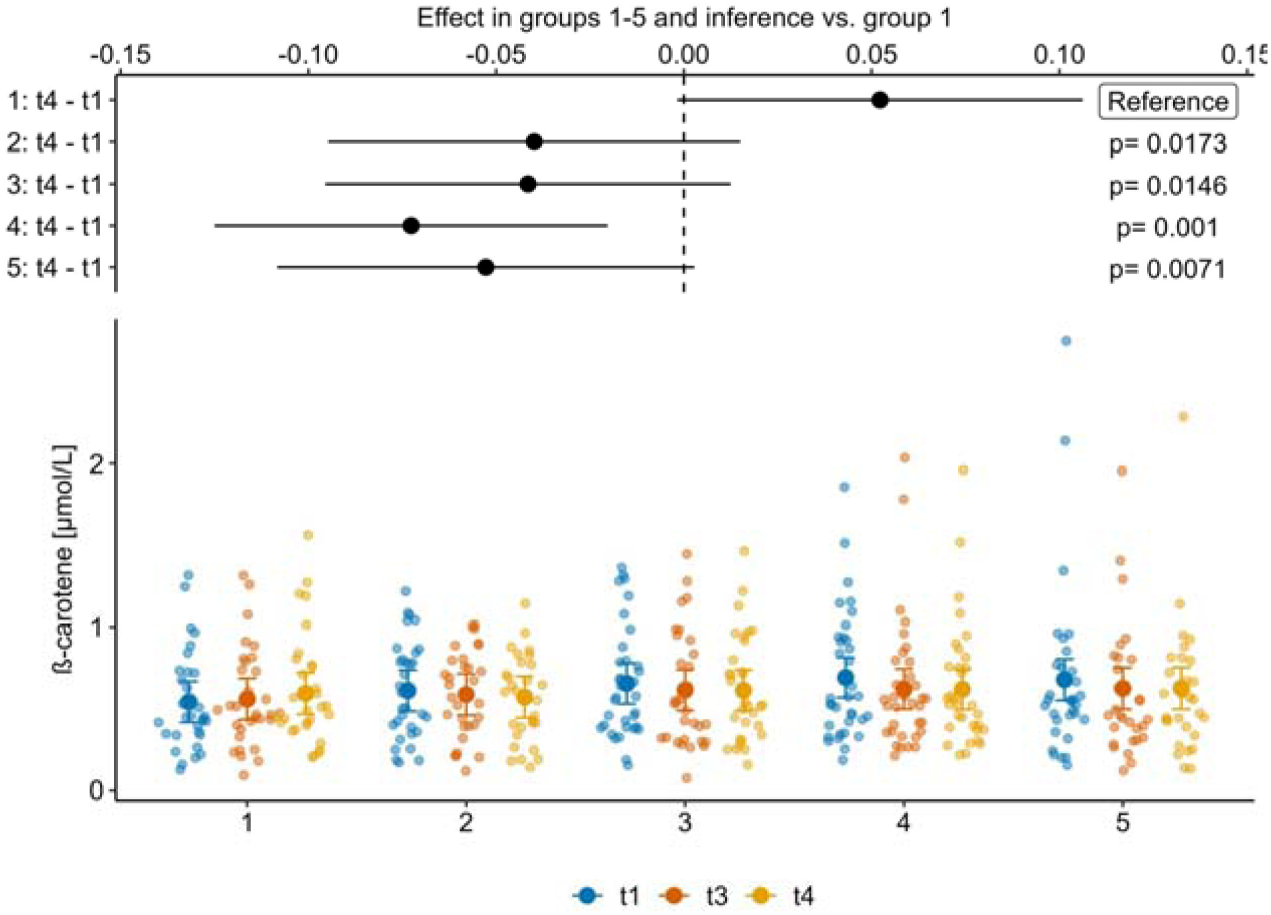
Effect plots and response data plots for selected secondary and for exploratory endpoints. (a) HDL; (b) sTNFr1; (c) GDF-15; (d) eGFR; (e) platelet counts; (f) β-carotene. See Figure 2 for further details.

**Suppl. Fig. 2.**
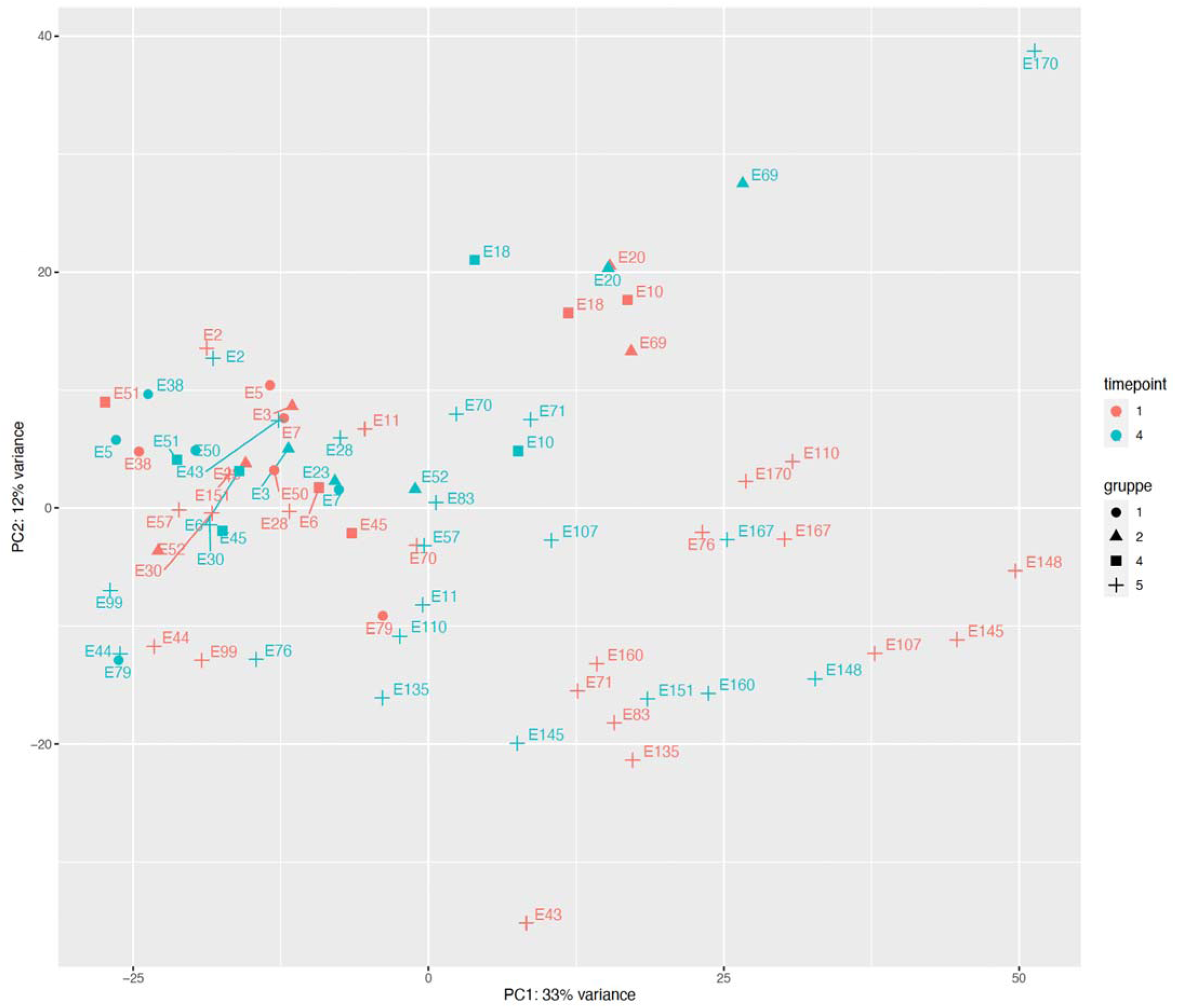
Principal Component Analysis of all NGS samples (taking into account only the top 500 variable genes across the samples). The color represents the time-point, the shape represents the group to which the samples belong.

