## Supplementary Methods Results and Discussion for "High-dose Polyphenol-rich Nutrition Improves Lipid and Inflammation Profiles and Triggers Apoptotic Signaling in Healthy Elderly People (the ErdBEHR Study)"

**Supplementary Text, Methods, Results and Discussion**

1) **Presentation of the group 1 gene expression data.**
2) **Box 1 Supplementary Results, Discussion and Methods.**
3) **Box 3 Supplementary Methods.**

***Supplementary Results and Discussion, gene expression analyses.***

***Presentation of the group 1 gene expression data.*** We also obtained gene expression data of 5 participants of the control group 1 for the first and the last time-point. As expected, longitudinally without an intervention, the signal in the data for group 1 is weaker than for the intervention groups; maximum q-values (on the negative log scale) of up to 0.35 are observed, compared to q-values up to 0.45 (in group 2; 5 participants profiled), up to 5 (in group 4; 5 participants profiled) and up to 14 (in group 5; 21 participants profiled). Nevertheless, similarly to the intervention groups, we find GOBP terms "enriched-up" related to mitochondria, immunity, cell-killing, metabolism and stress response (*leukocyte mediated cytotoxicity/ cell killing/ positive regulation of NF−kappaB/ ATP metabolic process*) (Fig. A). Similarly, the KEGG "enriched-up" pathways feature mitochondria, immunity and metabolism (*Natural killer cell mediated cytotoxicity/ Fc gamma R−mediated phagocytosis/ Oxidative phosphorylation/ Thermogenesis*) (Fig. B). The most likely explanation for these weak enrichments is a seasonal effect, given that participants were in the study during 10 weeks from mid-June to 1 October. In particular, immune challenges surface from the end of August onwards, as was also seen in seasonal patterns (Dopico et al., 2015) (their fig. 5a,c,d), reflecting in particular the seasonal upregulation of immune terms (incl. phagocytosis) and of genes (incl. CRP) from August onwards.

We refrain from interpreting the cross-sectional comparison between groups (e.g. of group 5 to group 1 at the (first or) last time point). Of note, a few of the 21 participants of group 5 contribute to high heterogeneity in gene expression patterns of group 5, in stark contrast to the cross-participant homogeneity of the 5 participants selected from group 1 (see, e.g., the heatmap for the transcripts underlying the enrichment of the GO term *phagocytosis* in the Suppl. Data); this heterogeneity does not cancel out in a cross-sectional comparison as it does in any longitudinal comparison where all participants serve as their own controls. See the Suppl. Data for the tables of DEGs, the enrichment data (tables), and more heatmaps.

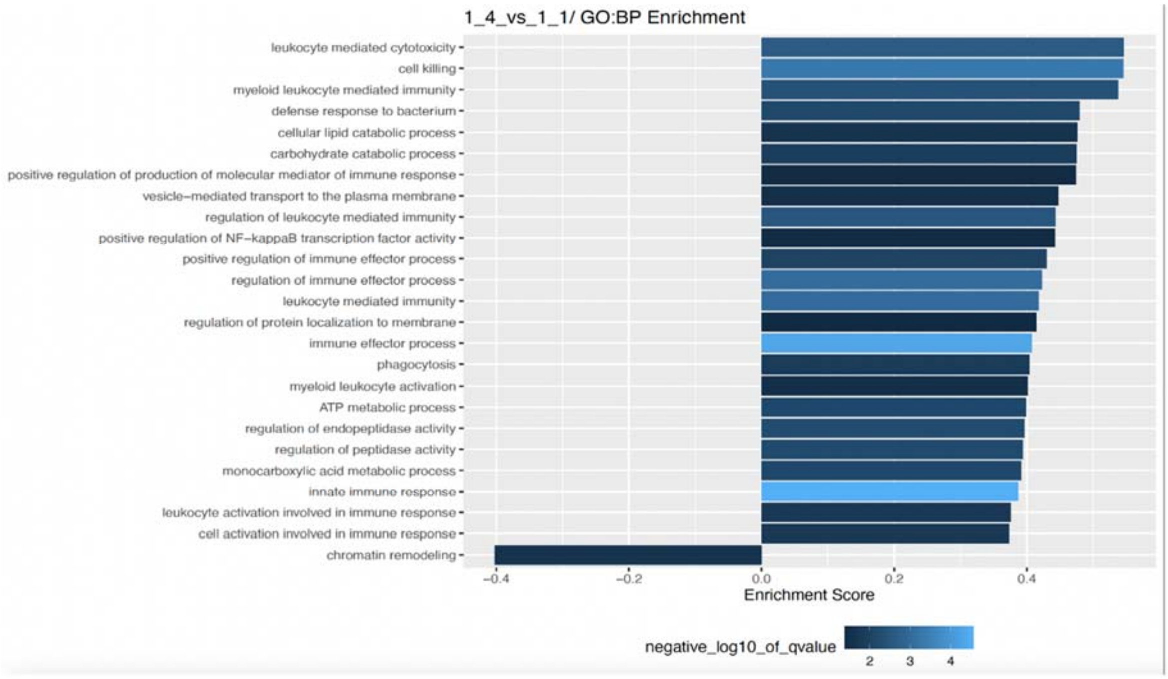


Fig. A. Differential Gene Expression enrichment results for group 1, and associated heatmap. Group 1 longitudinal differences t1 versus t4, ***enriched GOBP terms***. The size of the bar represents the *Enrichment score* and the color represents the negative log10 of the *qvalue* measuring the significance of the enrichment.

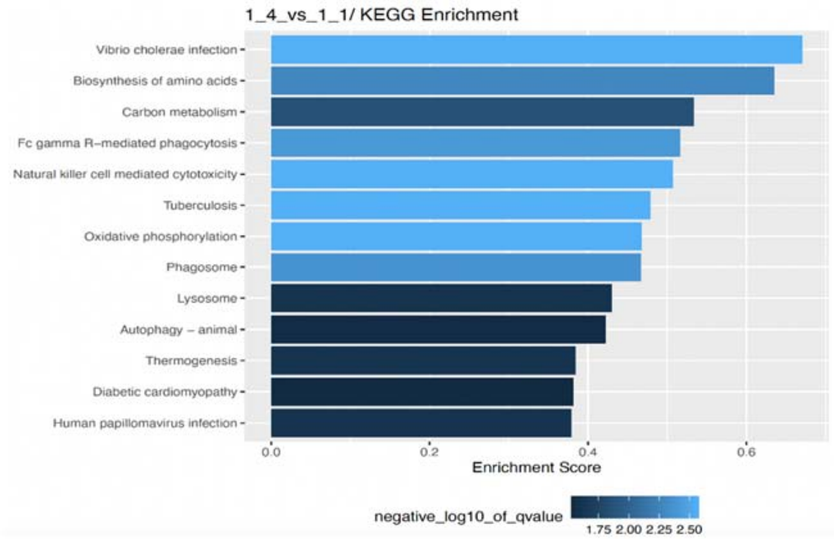


Fig. B. Differential Gene Expression enrichment results for group 1, and associated heatmap. Group 1 longitudinal differences t1 versus t4, enriched ***KEGG pathways***. The size of the bar represents the *Enrichment score* and the color represents the negative log10 of the *qvalue* measuring the significance of the enrichment.

### *Box 1 Supplementary Results, Discussion and Methods.*

***Mass spectrometric measurements of quercetin and fisetin in strawberries, capers, and controls.***

***Supplementary Results.***

Quercetin was detected in all strawberry varieties studied here. In particular, after treatment of the crushed strawberries with β-glucuronidase, a significant increase in free quercetin was detected after extraction of the samples, indicating that quercetin was previously strongly bound to glucuronides (Table A). Exemplary ion chromatograms for the MRM transitions of quercetin of a β-glucuronidase treated and an untreated strawberry sample are shown in Figure C. The increase was greatest in the second Florentina sample with approximately 55-fold increase, with the first Florentina sample showing the least increase with approximately 4-fold increase. As a positive control, samples of lovage or capers were included in each experiment, which showed a clear quercetin signal in the mass spectrometer. Here, significantly more quercetin was detected in lovage samples without β-glucuronidase treatment (148.4 µg/g) than with treatment (39.7 µg/g). Capers, on the other hand, showed a significant increase in concentration under β-glucuronidase treatment (108.4 µg/g vs. 46.1 µg/g in a freshly processed sample). Exemplary ion chromatograms for the MRM transitions of quercetin and acridine orange, as internal standard, of a β-glucuronidase treated and an untreated caper sample are shown in Figure D.

Table A: Measured amount of quercetin in strawberries. *n = 2*.

| Strawberry type | fresh extracted on the day of measurement (µg/g) | β-glucuronidase pretreatment, overnight incubation, extraction (µg/g) | β-glucuronidase pretreatment, overnight incubation, extraction (µg/g) |
|---|---|---|---|
| Malwina | 0.35 ± 0.18 | 0.43 ± 0.27 | 14.07 ± 4.20 |
| Florentina early August 2021 | 1.04 ± 0.45 | 0.43 ± 0.13 | 3.91 ± 0.64 |
| Florentina late August 2021 | 0.52 ± 0.06 | 0.26 ± 0.15 | 14.23 ± 4.74 |

In contrast to quercetin, fisetin could not be detected in any of the strawberry samples either freshly processed or after treatment with β-glucuronidase. In the positive control of smoketree samples included here, fisetin was detected both freshly worked up and after treatment with β-glucuronidase. Exemplary ion chromatograms for the MRM transitions of fisetin and acridine orange of a β-glucuronidase treated and an untreated smoketree sample are shown in Figure E.

Neither fisetin nor quercetin was detected in pilot-study-based serum samples tested at various time points (0;4;8;24 hours and 8 days). Treatment of the serum samples with β-glucuronidase also did not lead to any positive result.

***Supplementary Discussion.***

The strawberry cultivars examined in this study contained varying amounts of quercetin, regardless of cultivar. Most likely, quercetin concentration in strawberries is influenced by external factors such as the weather (sunlight or rain), or challenges such as insects or microbes. Quercetin was shown to protect the plants as an antioxidant and additionally strengthens various processes such as photosynthesis, growth and development (Singh, Arif, Bajguz, & Hayat, 2021). Most of the quercetin we could detect was accessible after treatment with β-glucuronidase. This is not surprising, since glucuronidation makes the substances more hydrophilic and they are then present in a dissolved form in the aqueous environment of the cell. Furthermore, it has already been found by other authors that quercetin is present as glucuronides in food (Boots, Haenen, & Bast, 2008). In particular, quercetin-3-O-glucuronide is known from wine (Satue-Gracia, Andres-Lacueva, Lamuela-Raventos, & Frankel, 1999), medicinal plants like *Hypericum hirsutum* or *Nelumbo nucifera* (Kitanov, 1988) (Kashiwada et al., 2005) and showed antimicrobial as well as antioxidant behaviour (Razavi, Zahri, Zarrini, Nazemiyeh, & Mohammadi, 2009).

On the other hand, fisetin was not detected in the strawberry cultivars studied here. Potentially, the failure to detect fisetin in our samples and the amounts found in the study of (Arai et al., 2000) can be explained by the different technology used at that time. Using our approach, employing various tests with different solvents and extraction variants, fisetin was not detected, using up to 50 g as input. In serum, fisetin was also not detected, though it was shown that fisetin is rapidly metabolised and thus may have eluded our analysis even if it was present in the food (Grynkiewicz & Demchuk, 2019).

Quercetin was not detected *in serum* above our detection limit (LOD 0.12 µM and LOQ 0.3 µM) even after treatment with β-glucuronidase. We cannot exclude that it was present especially in the first hours after strawberry ingestion. In a study with frozen onions, maximum concentrations were reached after about 3 hours and after 48 hours concentrations were very low (at 10 ng/ml) (Hollman et al., 1996). Furthermore, it was shown that in addition to quercetin, two monomethylated derivatives, isorhamnetin (3'-O-methyl quercetin), and tamarixetin (4'-O-methyl quercetin) can also predominate (Burak et al., 2017), but these were not considered in our study.

#### *Supplementary Methods.*

Several workup methods were used for the identification and quantification of fisetin and quercetin from strawberries, capers, and human plasma. For the former, the focus was on different varieties of strawberries (Malwina as well as Florentina from two different harvest dates) and capers in olive oil. As positive controls, samples of lovage (leaves, positive control quercetin) and smoketree (leaves and branches, positive control fisetin) were also examined. For analysis, 0.1-0.5 g of frozen mortared sample material was used. For the nearly anhydrous samples (capers, lovage, smoketree), 500 µl of water was added. This step was not necessary for the strawberries and was thus omitted. Subsequently, 50 µl of a 5 µM acridine orange solution was added to the samples as an internal standard. Ethyl acetate was chosen as the extraction solvent. The samples were then extracted twice with 1 ml ethyl acetate. After an extraction time of one hour, the samples were centrifuged at 14,000 rpm (26.342 g) for 5 min. The organic supernatant was concentrated to dryness in a vacuum concentrator (SpeedVac SPD130DLX, Thermo Fisher Scientific, Asheville, NC, USA) and then reconstituted in 100 µl of acetonitrile. This workup only captures free fisetin and quercetin and would not properly capture glucuronide-bound fisetin and quercetin. Therefore, additional samples to those described above were prepared one day before measurement to which β-glucuronidase was added. For this, 1 ml of 1 mM KH2PO4 buffer pH 4.5 and 25 µl of β-glucuronidase (≥ 1250 U/ml) from helix pomatia (Sigma Aldrich, Taufkirchen, Germany) were added to the sample material and incubated overnight at 37 °C with gentle shaking. Sample material without β-glucuronidase was also analyzed. Furthermore, 1 mM KH2PO4 buffer with and without the addition of β-glucuronidase was included as negative control. Before extraction on the following day, the internal standard was added as for the other samples. Subsequently, due to the larger initial volume of the samples with β-glucuronidase, 2 ml ethyl acetate was chosen twice for the extraction. After concentration to dryness, these samples were also reconstituted in 100 µl acetonitrile.

For analysis from human plasma, 500 µl of plasma was used for each of the three sample workups. Different time points (0;4;8;24 hours and 8 days) after strawberry and caper ingestion from five pilot study donors were analysed.

To assess the amount of substance in the sample material, a common serial standard row was prepared from stock solutions of fisetin (50 mM) and quercetin (60 mM), both dissolved in DMSO, in acetonitrile. A range of 0.05-0.5 mM was chosen for fisetin and 0.03-0.6 mM for quercetin. From these concentrations in acetonitrile, 10 µl was added to 990 µl of water followed by 50 µl of 5 µM acridine orange solution. The final concentration range was thus 0.5-5 µM for fisetin and 0.3-6 µM for quercetin. For the evaluation of plasma samples, the common serial dilution was performed directly in human plasma. Therefore, the final concentration range for fisetin was set to 0.25-5 µM and for quercetin 0.3-3 µM. Further sample processing is identical to the one described above.

5 µl from all described samples were injected for analysis by LC-MS/MS. Separation was achieved using a Shimadzu LC-20AD HPLC with a Multospher 120 C18 AQ column 125 × 2 mm, 5 µm particle size (CS-Chromatographie Service GmbH, Langerwehe, Germany) coupled to a guard column (20 mm × 3 mm , 5 µm particle size). Water was chosen as mobile phase A and acetonitrile as mobile phase B, both containing 0.2 % formic acid. The flow rate was set to 0.3 ml/min. First, the initial conditions of 15% mobile phase B were maintained for 3 min. Subsequently, a linear gradient up to 50 % B was selected for the next 7 min and then increased to 100 B within the next 2 min. Thereafter, the initial conditions were restored within 3 min and the column was re-equilibrated for another 5 min. The total run time for one sample measurement was 20 min. The oven temperature was set to 40 °C. Mass spectrometric analysis was carried out on a Shimadzu LCMS-8050 triple quadrupole mass spectrometer. Fisetin and quercetin were measured in positive and negative mode, while the internal standard acridine orange was measured only in the positive mode. The dwell time for all ions was set to 100 msec. For the assessment of the presence of fisetin and quercetin in the analysed samples all fragment ions and their relative intensity were used. The respective transitions and associated mass spectrometric settings as well as the general settings of the triple quadrupole are shown in Tables B/C.

Table B: Parameters of the triple quadrupole interface.

| Interface ESI parameter | Value |
|---|---|
| Nebulizing gas flow | 3 L/min |
| Heating gas flow | 10 L/min |
| Interface temperature | 300 °C |
| Desolvation temperature | 526 °C |
| DL temperature | 250 °C |
| Heat block temperature | 400 °C |
| Dry gas flow | 10 L/min |

Table C: Mass spectrometric parameters of fisetin, quercetin and the internal standard acridine orange.

| Substance | Precursor (*m/z*) | Product (*m/z*) | Q1 Pre Bias (V) | CE (V) | Q3 Pre Bias (V) | Measuring mode |
|---|---|---|---|---|---|---|
| Fisetin | 287.2 | 137.0 | -11 | -32 | -27 | positive |
| Fisetin | 287.2 | 213.0 | -10 | -29 | -22 | positive |
| Fisetin | 287.2 | 128.1 | -11 | -48 | -14 | positive |
| Fisetin | 285.0 | 135.0 | 20 | 22 | 13 | negative |
| Fisetin | 285.0 | 121.0 | 20 | 27 | 20 | negative |
| Fisetin | 285.0 | 163.15 | 20 | 17 | 16 | negative |
| Quercetin | 303.0 | 229.05 | -18 | -30 | -24 | positive |
| Quercetin | 303.0 | 152.9 | -12 | -33 | -30 | positive |
| Quercetin | 303.0 | 257.0 | -13 | -25 | -27 | positive |
| Quercetin | 301.0 | 151.2 | 21 | 21 | 14 | negative |
| Quercetin | 301.0 | 179.2 | 21 | 18 | 18 | negative |
| Quercetin | 301.0 | 106.95 | 21 | 28 | 22 | negative |
| Acridine Orange | 266.3 | 250.1 | -14 | -35 | -28 | positive |
| Acridine Orange | 266.3 | 234.0 | -14 | -52 | -26 | positive |
| Acridine Orange | 266.3 | 222.1 | -14 | -34 | -24 | positive |

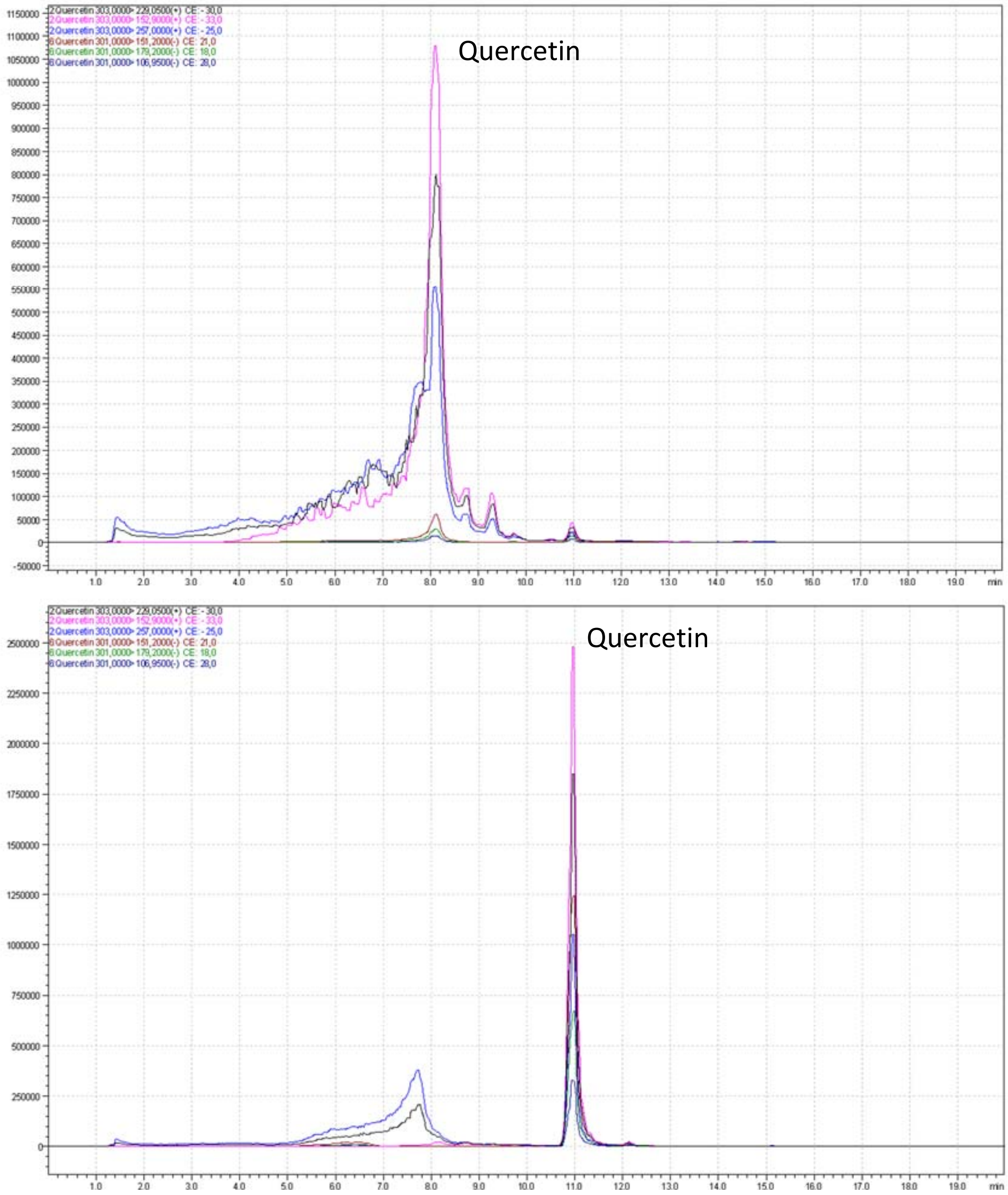


Figure C: Measured MRM transitions for quercetin of strawberry samples (Malwina). Shown are the mass traces from a freshly processed sample of Malwina (top) and a Malwina sample treated with β-glucuronidase (bottom).

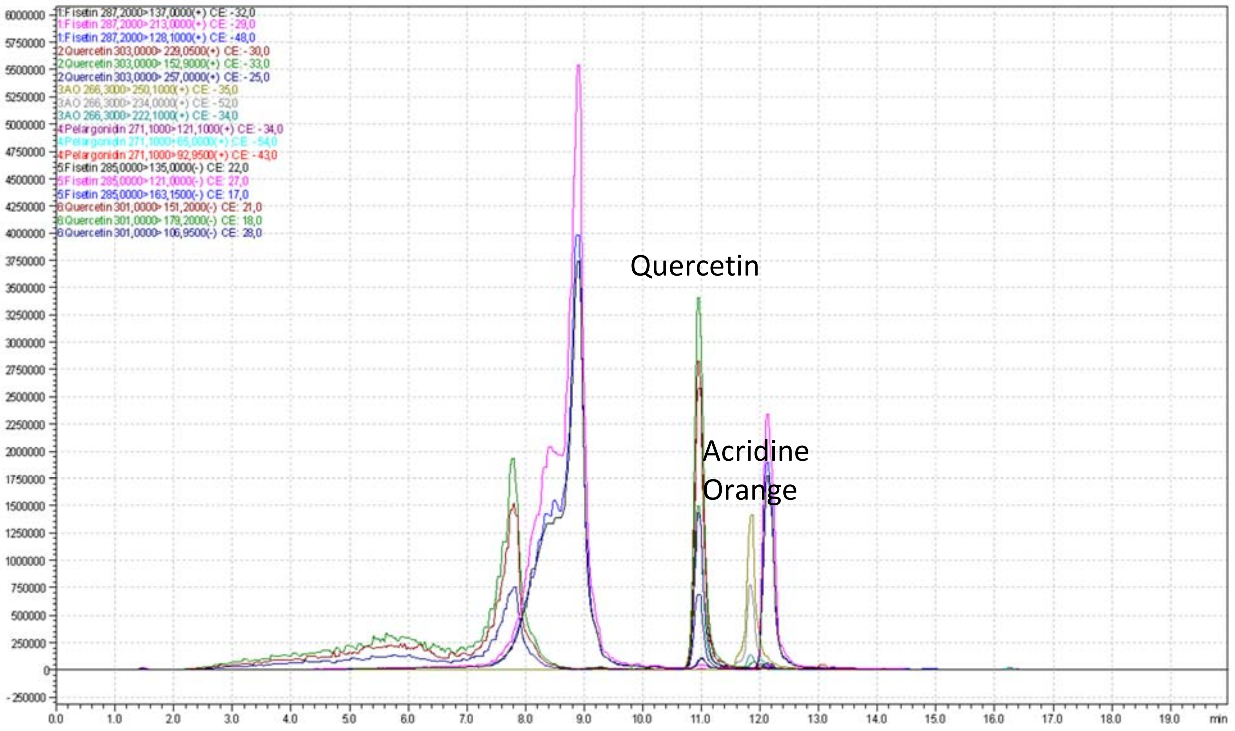


Figure D: Measured MRM transitions of a freshly processed caper sample.

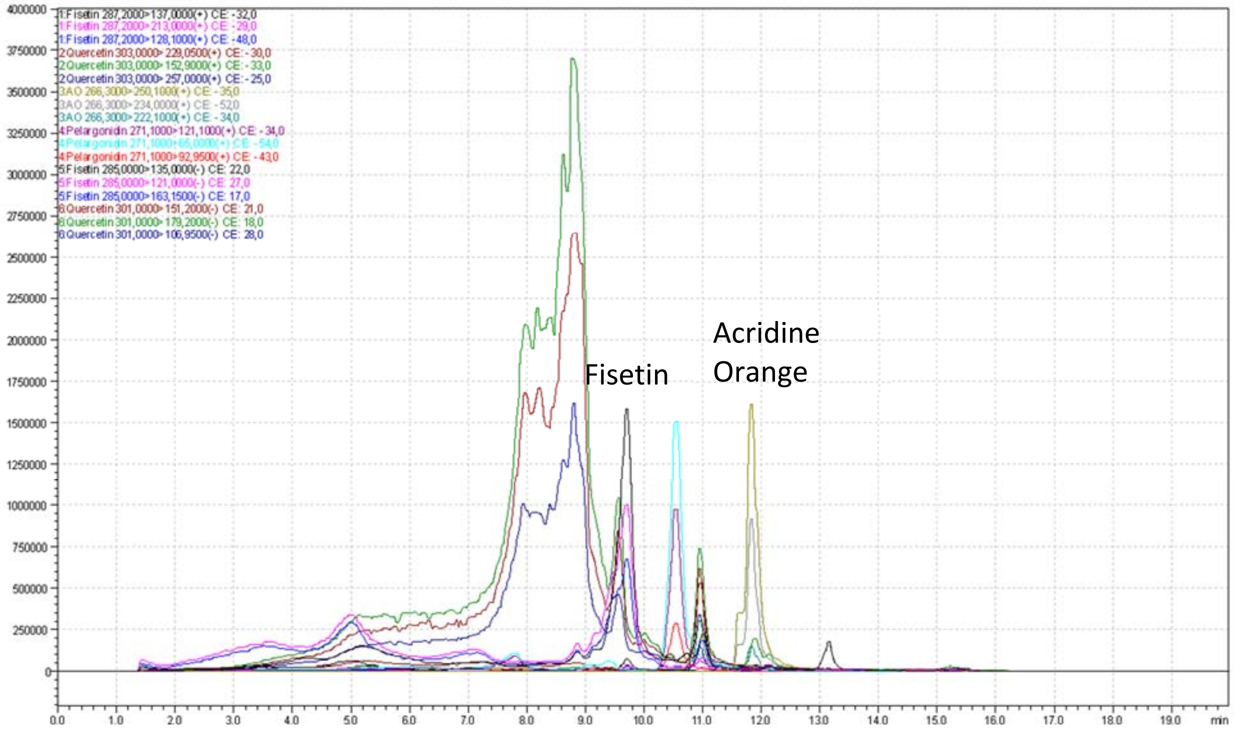


Figure E: Measured MRM transitions of a freshly processed smoketree sample.

***Box 3 Supplementary Methods.*** Induction of CRP in cultured human liver cells (Hep 3B) and measurement of *CRP* mRNA level were conducted as previously described {Lüersen, 2023, 10.3390/nu15061392} in accordance to others (Yoshida et al., 2006; Zhang, Jiang, Rzewnicki, Samols, & Kushner, 1995). Hep 3B cells were kindly gifted by Claudia Geismann (Laboratory of Molecular Gastroenterology & Hepatology, Department of Internal Medicine I, UKSH-Campus Kiel, 24105 Kiel, Germany). Cells were cultivated in MEM with Earle's balanced salt solution (EBSS), L-glutamine (PAN Biotech, Aidenbach, Germany) and 2.2 g/L NaHCO3, supplemented with 10% (vol/vol) heat inactivated fetal bovine serum (Gibco™ by Thermo Fisher Scientific GmbH, Life Technologies™, Darmstadt, Germany) as well as 1% penicillin/streptomycin (PAN Biotech, Aidenbach, Germany) for 5 days. For CRP induction, Hep 3B cells were incubated with a commercially available freeze-dried strawberry powder (myfruits ® Janzen Ventures GmbH, Kehl, Germany) resolved at 10 mg/ml in DPBS (PAN Biotech, Aidenbach, Germany). The supernatant obtained by short centrifugation of the resolved strawberry powder was subjected to sterile filtration. The cells were then incubated with the strawberry extract at indicated concentrations (from 5 µg/ml to 5 mg/ml) in serum-free media containing 1 µM of dexamethasone (Dex) and stimulated with interleukin-1ß (IL1ß, 400 U/ml) and interleukin-6 (IL6, 200 U/ml) (both from ImmunoTools GmbH, Friesoythe, Germany) for 18 h. DPBS served as solvent control at a final dilution of 50%. RNA isolation and quantitative RT-PCR was done as previously described (Lüersen et al., 2023), (Kuhn, Pallauf, Schulz, & Rimbach, 2018). In brief, cells were harvested and RNA isolated with peqGOLD TriFast (VWR International, Radnor, PA). Gene expression was analyzed via quantitative RT-PCR with the Sensi-FAST™ SYBR® No-ROX One-Step Kit (Bioline, Luckenwalde, Germany) using a Rotorgene 6000 cycler (Corbett Life Science, Sydney, Australia). Gene expression levels were determined using a standard curve and normalized to the expression level of *GAPDH*. Primers were as follows: *CRP* forward primer: 5'-CCCTGAACTTTCAGCCGAATACA-3'; *CRP* reverse primer 5'-CGTCCTGCTGCCAGTGATACA-3'; *GAPDH* forward primer: 5'-CAATGACCCCTTCATTGACC-3'; *GAPDH* reverse primer: 5'-GATCTCGCTCCTGGAAGATG-3'. Statistical analysis was performed with the software GraphPad Prism (Ver. 7.05) comparing the different strawberry extract concentrations with the solvent control DPBS in a Kruskal-Wallis ANOVA with an uncorrected Dunn's test. A p-value <0.05 was considered significant.
